# Association of cognitive impairment with statin use in coronary artery disease across APO (ε) genotypes in AllofUS

**DOI:** 10.64898/2026.06.02.26354765

**Authors:** Praveen Hariharan, Minoo Bagheri, Frank Sellke

**Affiliations:** Warren Alpert Medical School, Brown University, Providence, RI; Vanderbilt University Medical Center, Nashville, TN

**Keywords:** Dementia, Mild Cognitive Impairment, APO (ε) genotype, Statin use, Coronary artery disease

## Abstract

**BACKGROUND:** Coronary artery disease (CAD) and Impaired Cognitive (IC) disease share sociodemographic, genetic, and clinical factors, but the association of IC with statin use in CAD remains unclear.

**OBJECTIVES:** To determine the association between IC and statin use in CAD based on APO (ε) genotype, sex, and lipid levels.

**DESIGN, SETTING, AND PARTICIPANTS:** We performed a retrospective study of AllofUS (AoU) participants with CAD (age≥60 yrs) enrolled from 2017 to 2023. We defined CAD as having a history of angina/myocardial infarction/chronic ischemic heart disease or having percutaneous coronary intervention/CABG, and IC defined as mild cognitive impairment or all-cause dementia, using ICD/SNOMED codes.

**MEASURES:** We assessed the association between IC and statin use using logistic regression analysis, while adjusting for clinical factors, sociodemographics, and APO (ε) genotypes before and after propensity score matching. We further performed stratified analysis by sex, and APO (ε) genotypes. We finally assessed the association between IC and statin users, based magnitude on the change in lipid levels before CAD and after IC (TC-Total cholesterol, LDL – low density lipoprotein, HDL-High Density Lipoprotein). Significance was defined at p < 0.05.

**RESULTS:** The cohort included 22,089 participants with CAD and 1343 with IC. Thirty-nine percent of participants were females, 77% were European, 13% were African American, and 9% were of Admixed American ancestry. The proportion of IC was higher (**6.8% vs 3.5%, p<0.001**) in statin users (n=17,191) vs non-statin users (n=4,898). IC was significantly associated with statin use (**OR:1.70;1.40-2.10, p = 4.9e-7**) after adjustment for clinical factors, sociodemographics, and APO (ε) genotypes. After propensity-score matching between IC and CAD, we observed an association between IC and statin use (**OR:1.55;1.24-1.94, p =1e-4**). In stratified analysis, the association between IC and statin use was strongest in the APO ε3/ε3 group (**OR:2.04;1.53-2.75, p = 1e-6**), and in females (**OR:2.20;1.60-3.06, p = 2.e-6**) compared to males (OR:1.43;1.10-1.90, p = 0.01). We finally observed an increased magnitude of association between IC and statin users having higher HDL increase (**> 10 mg/dl: OR:1.95;1.44-2.66, p=1e-5**) as compared to statin users with lesser HDL increase (**≤ 10mg/dl: OR:1.61;1.22-2.15, p=8e-4**).

**CONCLUSION:** In the AllofUS cohort, IC was significantly associated with statin use in CAD participants. We observed the strongest association in the APO ε3ε3 group, among females, and with a greater increase in HDL levels in statin users.

**Key Message:** *What is already known on this topic:* Coronary artery disease (CAD) and Impaired Cognitive (IC) disease, i.e., mild cognitive impairment or all-cause dementia, share genetic, sociodemographic, and clinical risk factors but the association of IC with statin use in CAD remains unclear.

*What this study adds:* We observed an association between IC and statin use in CAD participants after adjusting for sociodemographics, clinical factors, and APO (ε) genotypes. The association persisted after propensity score matching for sociodemographics, clinical factors, and APO (ε) genotypes. Further, when CAD participants were stratified across APO (ε) groups and by sex, we observed strongest magnitude of association between IC and statin use in the APO ε3ε3 compared to other APO (ε) genotypes, and in females compared to males. Among CAD participants with documented baseline and latest lipids (total cholesterol (TC), low-density lipoprotein (LDL), high-density lipoprotein (HDL)), IC was associated with statin use regardless of baseline lipid levels or latest lipid levels. While we did not observe any change in association between IC and statin use, based on the magnitude of decrease in TC and LDL levels, we observed a higher magnitude of association between IC and statin use, with a greater increase in HDL levels.

*How this study might affect research, practice or policy:* Our observations highlight the association of IC and statin use in CAD and the role of APO (ε) genotype evaluation, and serial lipid level assessments for evaluating for statin associated IC in CAD.

## INTRODUCTION

Age associated Impaired Cognitive Disease (IC), i.e, all-cause dementia (ACD) or mild cognitive impairment (MCI), and Coronary Artery Disease (CAD) are a significant cause of morbidity in the aging societies, with associated increased financial burden to health care systems and patients.^1^ IC after a CAD event is a matter of concern and its prevalence increases in the geriatric group (≥60 years).^2,3^ Further IC as a clinical entity may have heterogeneous neuropathologies, yet traditional clinical factors like age, sex, diabetes, hypertension, hyperlipidemia, depression, ischemic stroke, obstructive sleep apnea, chronic kidney disease, smoking, alcohol use, educational background, APO (ε) genotypes, are shared by CAD and IC patients.^1,3,4^

Statins are widely used for primary and secondary prevention of CAD, yet their association with IC has been conflicting.^5–12^ Further, in the advanced geriatric group (>75 years), the utility of statin for CAD prevention outside of high-risk cases has been debated, with no randomized trials supporting statin use for CAD risk reduction.^5,13^ While considering the association between IC and statin use in CAD patients, factoring in APO (ε) genotypes, sex, and lipid levels can help in dissecting the association more clearly.^8,14–19^ IC and CAD share underlying genetic architecture related to dyslipidemia, in particular APO (ε) genotypes have been identified to be jointly associated with CAD and IC, and can be an effect modifier when assessing the association of statin use with IC in CAD.^8,20–23^ A recent study of evaluated the association between cholesteryl ester transfer protein inhibitor (CETPi) and changes in neurodegenerative biomarkers (ptau-217, AB42/40, GFAP, NfL) across APO (ε) genotypes in atherosclerotic cardiovascular disease patients on high dose statins after 1 year of CETPi randomization.^25^ The investigators noted an increase in ptau-217 levels across most APO (ε) genotypes groups, and more so in the statin treated placebo group, except for reduction in ptau-217 levels in APO ε4ε4 group receiving CETPi. The increase of ptau-217 was more so in the ε4ε4, ε3ε4, ε2ε3 groups followed by ε3ε3 groups with similar patterns noted across other neurodegenerative biomarkers. These observations suggest variable interaction between lipid-modifying agents and neurodegenerative biomarkers based on APO (ε) genotype and proves the association of lipid modifying agents including statins in neurodegenerative disease pathophysiology. Further in a large UK biobank study of dementia free individuals at baseline (age >40 yrs), statin users had higher hazard ratio (HR) for incident Alzheimer’s Disease (AD) compared to non-statin users.^8^ In addition, through drug target Mendelian randomization (MR), HMGCR single-nucleotide variants (SNV) corresponding to low LDL-C levels were associated with lower cognitive performance and brain cortical surface area.^24^ Also, statin users had higher HR for AD in females as compared to males in the general UK biobank cohort.^8^ Finally, extreme lipid level fluctuations in the setting of medication use and/or lifestyle factors have also been associated with IC in geriatric group.^18,19^ In this context, large studies in CAD participants evaluating the association of IC and statin use while accounting for APO (ε) genotypes, and assessing effect modification related to sex or APO (ε) genotypes are lacking. Given that lipid level variations go hand in hand with statin use, it is unclear if the association between IC and statin use varies based on lipid level variations in CAD.^10,15,18^ Understanding the association between IC and statin use in CAD patients in this context would provide physicians with tools for statin use in the geriatric CAD population.

In this study, we sought to evaluate the association between IC and statin use in all CAD participants with consenting age ≥ 60 years at the time of enrollment in the diverse NIH AllofUS (AoU) cohort enrolled between 2017 and 2023. We further dissected the association between IC and statin use across individual APO (ε) genotype groups, and sex. Finally, we assessed the association between IC and statin use based on lipid level variations in CAD participants.

## METHODS

### a) Study Design

**Figure 1** highlights the retrospective design. We report our study according to STROBE guidelines.

**Figure 1:**
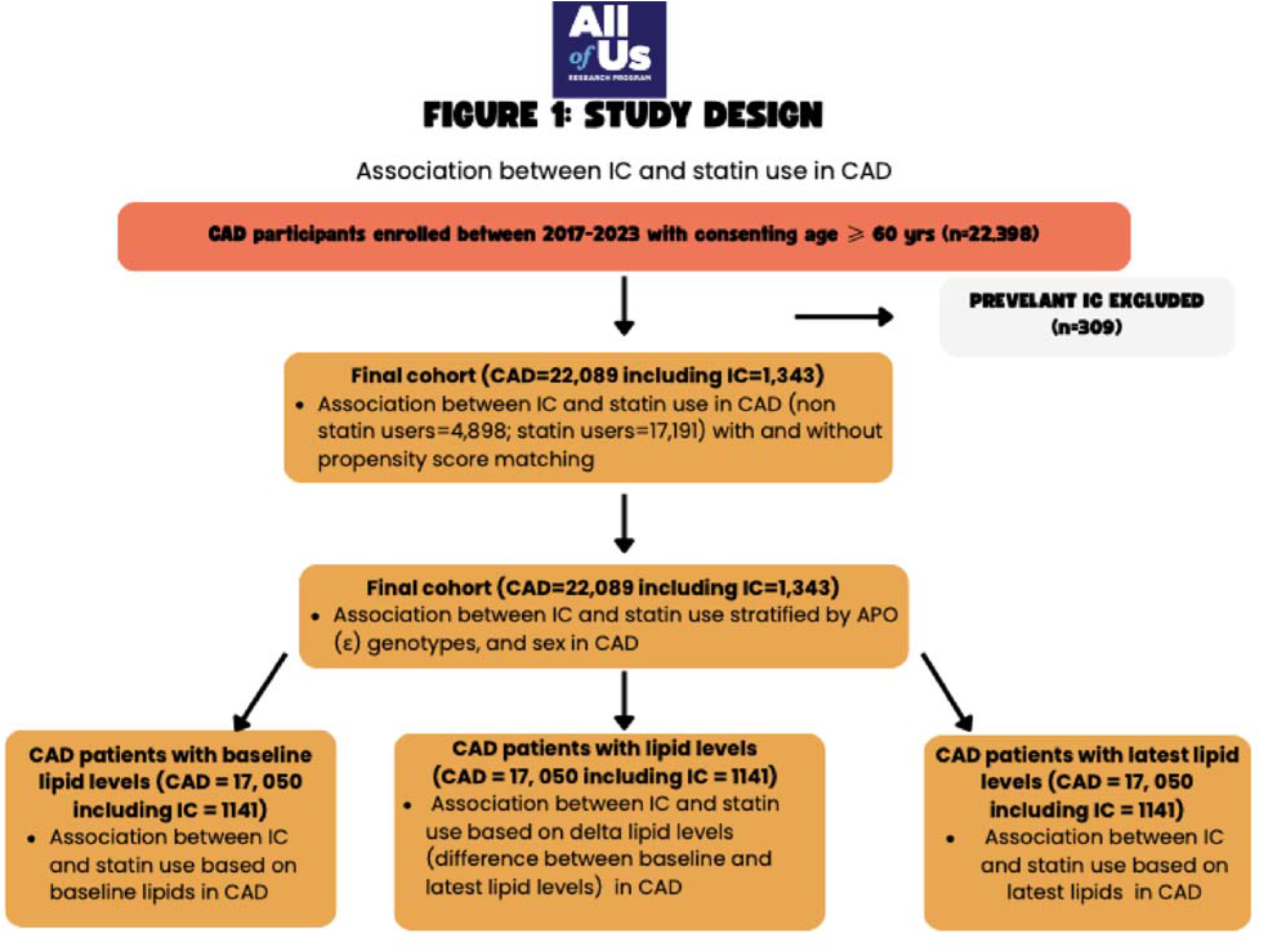
IC: Impaired cognitive disease defined as all-cause dementia or mild cognitive impairment CAD: Coronary artery disease

### b) Setting and Data Sources

In brief, the AoU prospectively assembles diverse participants across 50 healthcare organizations in the US and has gathered surveys, electronic health records (EHR), biosamples, and genomic data since 2017.^26^ AoU utilizes the Observational Medical Outcomes Partnership Common Data Model (OMOP CDM) based on SNOMED (Systematized Nomenclature of Medicine), and/or ICD 9/10 codes to standardize data.^26^ We used the AoU version 8 (AoU, v8) dataset (C2024Q3R8, released 2025) containing EHR data of more than 393,596 patients up until 2023.^27^

### c) Selection of Cohort

We used OMOP CDM v 5.3.1 concept codes and names, which are based on SNOMED and ICD-9/10 codes to define CAD, IC, covariates, and used the associated index date of diagnoses (Details in Supplement 1).^28–31^

#### CAD definition

Any participant with 2 or more visit encounters meeting one of the following criteria: A history of myocardial infarction (MI) (ICD-9: 410.x-412.x; ICD-10: I21.X-I24.X); the presence of stable or unstable angina (ICD-9: 413.x; ICD-10: I20.X); a history of chronic ischemic heart disease (ICD-9:414.x; ICD-10: I25.X); a history of percutaneous coronary intervention (PCI, ICD-10: Z95.5, Z98.61) or CABG (ICD-10: Z95.1).^22,28,32^ We also recorded the index date of CAD diagnosis.

#### Primary outcome

We have defined IC when participants have 2 or more encounters meeting the ICD-9/10 diagnosis for all-cause dementia or mild cognitive impairment (290.X, 294.X, 331.X, F01.X, F02.X, F03.X, G30.X, G31.X).^28,29,33,34^ All events were captured via the EHR up until 2023. We defined IC to include the entire spectrum of cognitive impairment, required a minimum age of 50 years at the time of IC diagnosis, and also recorded the index date of IC diagnosis. We further excluded IC before CAD diagnosis to remove prevalent IC cases. The inclusion of age cut-offs and ICD codes has been shown to achieve good accuracy in defining IC in large registries.^29,34^

#### Secondary Outcome

To assess the association between statin use and neurocognitive test skills in CAD participants, we used the quantitative gradual onset continuous performance task (GradCPT) accuracy response (d-prime), collected using unsupervised Explore the Mind (EtM) surveys from AoU participants in 2023 (C2024Q3R5, released Feb 2025).^27^ GradCPT assesses core executive function by testing sustained attention and inhibitory control.^35^

### d) Predictors

#### 1.1) Statin use

We defined statin prescription use based on OMOP concept ID (21601855), which is the parent concept ID for all HMG-CoA reductase inhibitors. (Details in Supplement 1).

#### 1.2) Lipid level data

For accessing lipid level data, we used the AoU, v8 data set to extract all total cholesterol (TC), low-density lipoprotein (LDL), and high-density lipoprotein (HDL) measurements based on the corresponding concept ID (Details in Supplement 1) and recorded the earliest lipid levels (baseline lipids) and the most recent lipid levels (latest lipids), and calculated the magnitude of change in lipid levels separately. We finally restricted all participants with lipid levels to only our CAD participant list.

#### 1.3) Systolic Blood Pressure (SBP) data

We obtained SBP data using concept ID (Details in Supplement 1) and restricted it to participant-provided information collected at the time of enrollment. We recorded the lowest SBP when multiple measures were recorded at enrollment.

#### 1.4) Clinical covariates

We further defined our covariates based on survey data and OMOP standard concept names and codes, which were based on SNOMED/ICD 9/10codes (Details in Supplement 1), and also recorded the index date of diagnoses. ^4,28,30,31^

##### a) Demographics

Age, self-reported sex, and principal component computed genetic ancestry.^26^

##### b) Social-determinants

We obtained the highest educational status, health insurance information, annual household income, current employment status, and community deprivation index (CDI: based on the zipcode of the participant) from baseline survey data received at the time of enrollment. In brief, CDI is based on a composite of six census tract-level variables obtained from the 2017 American Community Survey that includes poverty fraction, median household income, fraction of high-school graduation rates among 25 years and older, fraction of population with health insurance, fraction of households receiving public assistance income/food stamps, and fraction of vacant houses. The CDI ranges from 0-1, with higher values indicating greater deprivation, and we subsequently categorized CDI into low, medium, and high tertiles for the purpose of our analysis (Details in Supplement 1).^36,37^

##### c) Traditional clinical factors

Hypertension, diabetes, hyperlipidemia, ischemic stroke, history of depression, obstructive sleep apnea (OSA), chronic kidney disease (CKD), antihypertensive prescription data (betablockers, angiotensin receptor blockers, calcium channel blockers, diuretics), antidiabetic prescription data (insulin, biguanides, sulfonylurea, thiazolidinediones, alpha glucosidase inhibitors, dipeptidyl peptidase 4 inhibitors, glucagon like peptide-1 inhibitors, sodium-glucose-cotransport 2 inhibitors), smoking, alcohol use, and Body Mass Index (BMI) data were obtained based on OMOP standard concept names and codes (Details in Supplement 1).^30,38^

##### b) Genotype covariates

We defined the APO (ε) genotype based on rs429358 (T/C) and rs7412 (C/T) single-nucleotide variants (SNV).^23^ For this analysis, we used the exon-joint call set with 45,704,594 variants across 414,797 individuals (AoU, version 8). We subsequently filtered the joint call set to include only our unique CAD participants. We further removed related samples and performed sample and variant-level QC with a call rate >95%. We further grouped APO (ε) into ε2ε2 (rs429358:TT; rs7412:TT), ε2ε3 (rs429358:TT; rs7412:CT), ε3ε3 (rs429358:TT; rs7412:CC), ε2ε4 (rs429358:CT; rs7412:CT), ε3ε4 (rs429358:CT; rs7412:CC), ε4ε4 (rs429358:CC; rs7412:CC) variant groups based on rs429358, rs7412 genotypes.^23^ Given that ε3ε3 has the most common genotype frequency, we used ε3/ε3 as the reference group for our primary logistic regression analysis.^23^

### e) Standard Protocol Approvals, Registrations, and Patient Consents

Brown University Research Agreements and Contracting Committee reviewed our retrospective analysis proposal and approved the use of the AoU researcher workbench. In addition, we have reviewed and completed the AoU data access requirements and are in compliance with the data user code of conduct for registered and controlled-tier data. No humans or animals are involved in this retrospective analysis.

### f) Statistical Analysis

#### 1) Primary Analyses

##### 1.1) Statin use analysis

For our primary analyses, we included participants who met the CAD definition and had age ≥ 60 years at the time of enrollment. We subsequently queried for IC diagnoses in CAD participants and grouped participants into IC (with CAD) and CAD (without IC). We analyzed baseline characteristics across IC and CAD participants using simple means and proportions. We further compared the proportions of IC across all CAD participants with and without statin use. We also compared the proportions of IC with and without statin use, stratified by APO (ε) genotypes and sex. For univariate analysis, we used the chi-squared test. To assess association between IC and statin use in CAD participants, we performed logistic regression for IC as dependent variable and statin use as independent variable, while adjusting for demographics (age, sex, ancestry), traditional clinical factors (coronary artery bypass graft, diabetes, hypertension, hyperlipidemia, depression, ischemic stroke, obesity (BMI ≥ 30), obstructive sleep apnea, chronic kidney disease, smoking, alcohol use, antihypertensive use, antidiabetic use), social determinants (health insurance, education (college graduate and/or advanced degree), annual household income, employment status, and community deprivation index), and APO (ε) genotypes. Subsequently, we assessed the association between IC and statin use by running similar logistic regression models stratifying CAD participants by APO (ε) genotypes and sex. We defined significance at p < 0.05 for this analysis.

##### 1.2) CAD cohort with lipid levels and SBP (Details in Supplement 1)

Given that changes in lipid levels go hand in hand with statin use, which can be influenced by APO (ε) genotypes, we wanted to explore the joint effect of statin use and lipid levels with IC in CAD and across APO (ε) genotypes.^15^ For this analysis, we performed the series of steps to extract lipid level data, and categorize lipid levels (Details in Supplement 1). We categorized baseline and latest lipids as TC130 (≥ 130mg/dl or < 130 mg/dl), HDL40 (≥ 40mg/dl or < 40mg/dl) and LDL (≥55 mg/dl, or <55 mg/dl). We used these categories to somewhat mirror lipid targets used for secondary CAD prevention and to assess joint association of statin and extreme low lipid targets with IC.^5,18^ Finally, we quantified the magnitude of difference between baseline lipids (first TC, HDL, and LDL) and latest lipids (latest TC, HDL, and LDL) as delta TC, delta HDL, and delta LDL, respectively. We categorized delta lipids as deltaTC50 (> 50 mg/dl, reduction), deltaHDL10 (>10 mg/dl, increase), and deltaLDL30 (>30 mg/dl, reduction). We compared baseline and latest lipids between statin and non-statin users in all CAD participants, and stratified by APO (ε) genotypes and sex. We subsequently, assessed the association between IC and statin use based on delta lipids using joint effect terms (statin: deltaTC50, statin: HDL10, and statin: deltaLDL30).

#### 2) Sensitivity Analysis (Details in Supplement 1)

We performed the following sensitivity analyses to ensure consistency of our findings:

##### a) Propensity-score matched analyses

Given that the traditional clinical factors in IC can confound the association of between IC and statin use and CAD, we propensity-matched the IC (with CAD) to CAD (without IC) for demographics, traditional clinical factors, social determinants, and APO (ε) genotypes and repeated our regression analysis to assess the association of between IC and statin use. b) Duration of statin use:

To assess the association between IC and duration of statin use in CAD, we calculated the total years of statin use for each CAD participant as the difference between the index year of statin therapy recorded in the EHR and 2023 (when the EHR data were released). Subsequently, we categorized each CAD participant as having used statins for >8 years or ≤8 years and assessed association between IC and duration of statin use (> 8 years vs ≤ 8 years) by running similar regression analysis. c) Time to event analysis:

In order to account for event time distribution, we further included only IC cases after the study enrollment and performed a Cox-proportional hazards model to estimate the hazard ratio (HR) for statin use on IC risk, while using consenting age and current age as two time points in the model and adjusted for clinical factors, social determinants, and APO (ε) genotypes.

##### d) EtM survey data analysis

Given the heterogeneity of EHR-based IC definitions, we assessed the association between neurocognitive test scores (GradCPT) and duration of statin use using linear regression.

#### 3) Statistical tools

We performed all analyses in R, integrated within Jupyter Notebooks (a secure, access-controlled, cloud-based analysis environment) on the AoU researcher workbench. We used the matchit and samplesizeestimator packages for propensity-score matching and sample size estimation.^39^ We further used survival package for time to event analysis. For primary analyses, we estimated 3.5% probability of IC in the non-exposed group, and to detect a minimum odds ratio (OR) of 1.5 among statin users (exposed), we calculated ∼2300 statin user cases (1:1 exposed: unexposed ratio) to be required for achieving 80% power at an alpha level of 5%.

## RESULTS

We included 22,089 CAD participants (IC with CAD = 1,343; CAD without IC = 20,746) in our final analysis (**Figure 1**).

### 1) Baseline characteristics

When we compared baseline characteristics (**Table 1**), we noted that participants with IC were significantly older (80.07 years vs 76.69 years, p<0.001) than those with CAD alone. The average age of onset of CAD was 66.7 yrs (± 8.5 yrs) and average age of onset of IC was 74.7 yrs (± 8 yrs). When we compared ancestries, we observed higher proportions of African Americans (14.6% vs 12.5%, p=0.02) and Admixed American (10.7% vs 8.4%, p=0.003) in IC than in CAD alone. IC had a significantly higher proportion of traditional clinical factors than CAD alone. IC had higher proportions of APO ε3ε4 (29.0% vs 21.2%, p < 0.001) and APO ε4ε4 genotypes (3.6% vs 1.9%, p < 0.001) than CAD alone. We noted a mean EHR data length of 15 years (IQR: 8-21 years).

**Table 1:** Baseline characteristics (AllofUS, v8, Age ≥60)

|  | CAD without IC (n=20,746) | IC with CAD (n=1,343) | P value |
| --- | --- | --- | --- |
| **Demographics\*** |  |  |  |
| Age (mean, SD) | 76.69 (62-105, SD=7.1) | 80.07 (64-105, SD=7.4) | **p<0.001** |
| Female sex | 8,022 (38.7%) | 502 (37.4%) | p=0.36 |
| **Genetic ancestry** |  |  |  |
| Europeans | 16,024 (77.2%) | 979 (72.9%) | **p<0.001** |
| Africans | 2,586 (12.5%) | 196 (14.6%) | **p=0.02** |
| Admixed Americans | 1,738 (8.4%) | 144 (10.7%) | **p=0.003** |
| Others | 398 (1.9%) | 24 (1.7%) | **p=0.82** |
| **Traditional clinical factors** |  |  |  |
| Depression | 7,731 (37.3%) | 874 (65.1%) | **p<0.001** |
| Ischemic Stroke | 2,526 (12.2%) | 384 (28.6%) | **p<0.001** |
| Hyperlipidemia | 19,330 (93.2%) | 1,301 (96.9%) | **p<0.001** |
| Hypertension | 18,867 (90.9%) | 1,299 (96.7%) | **p<0.001** |
| Diabetes | 9,924 (47.8%) | 802 (59.7%) | **p<0.001** |
| Obesity | 9,161 (44.2%) | 570 (42.4%) | p=0.220 |
| Highest Education | 10,086 (48.6%) | 571 (42.5%) | **p<0.001** |
| Statin use | 16,019 (77.2%) | 1,172 (87.3%) | **p<0.001** |
| Antihypertensive use | 17,980 (86.7%) | 1,239 (92.3%) | **p<0.001** |
| Antidiabetic use | 9,796 (47.2%) | 750 (55.8%) | **p<0.001** |
| CABG | 7,734 (37.3%) | 621 (46.2%) | **p<0.001** |
| Obstructive sleep apnea | 7,556 (36.4%) | 652 (48.5%) | **p<0.001** |
| Chronic kidney disease | 7,294 (35.2%) | 668 (49.7%) | **p<0.001** |
| **Social determinants** |  |  |  |
| Health insurance | 20,141 (97.1%) | 1,294 (96.4%) | p=0.248 |
| Smoking | 4,734 (22.8%) | 343 (25.5%) | **p=0.022** |
| Alcohol Use | 1,783 (8.6%) | 169 (12.6%) | **p<0.001** |
| CDI (Low)† | 8,282 (39.9%) | 430 (32.0%) | **p<0.001** |
| CDI (Medium/High)† | 11,780 (56.7%) | 873 (65%) | **p<0.001** |
| Annual Income > \$75,000 | 6,591 (31.8%) | 280 (20.8%) | **p<0.001** |
| Employment (Retired) | 13,327 (64.2%) | 965 (71.9%) | **p<0.001** |
| **APOE4 (rs429358) genotype** |  |  |  |
| TT | 15,509 (74.8%) | 869 (64.7%) | **p<0.001** |
| CT/CC | 5,237 (25.2%) | 474 (35.2%) | **p<0.001** |
| **APO (ε) genotypes** |  |  |  |
| **APOε2ε2** | 133 (0.6%) | 7 (0.5%) | p=0.72 |
| **APOε2ε3** | 2,451 (11.8%) | 131 (9.8%) | **p=0.02** |
| **APOε3ε3** | 12,925 (62.3%) | 731 (54.4%) | **p<0.001** |
| **APOε2ε4** | 445 (2.1%) | 35 (2.6%) | p=0.30 |
| **APOε3ε4** | 4,404 (21.2%) | 390 (29.0%) | **p<0.001** |
| **APOε4ε4** | 388 (1.9%) | 49 (3.6%) | **p<0.001** |

\*Proportions in bold with P value <0.05 (Pearson's Chi-squared test, baseline group = CAD)
Obesity (Body mass index ≥ 30)
Highest Education: College graduate and/or advanced degree
CAD: Coronary Artery Disease; IC: Impaired Cognitive Disease (defined as all-cause dementia or mild cognitive impairment);
CABG: Coronary Artery Bypass Graft
†CDI: Community Deprivation index. CDI not reported in (n=724) participants

### 2) Association of statin use with IC in all CAD participants (n=22,089)

Among all CAD participants, we observed that the proportion of IC was significantly higher in statin users (**6.8% vs 3.5%, p<0.001**) than in non-statin users (**Table 1, Supplement**). We also noted higher proportions of all types of IC among statin users than among non-statin users in CAD (**Table2, Supplement 1**). When we stratified participants across APO (ε) genotypes, we observed a high proportion of IC with statin use in APO ε3ε3 (**6.1% vs 2.8%, p<0.001**) and APO ε3ε4 (**9.1% vs 4.5%, p<0.001**) groups (**Table 1, Supplement**). We also observed higher proportions of IC in both male statin users (**6.7% vs 3.9%, p<0.001**) and female statin users (**6.9% vs 2.9%, p<0.001**) as compared to non-statin users (**Table 1, Supplement**).

In the multivariable logistic regression model after adjusting for demographics, traditional risks, social determinants, and APO (ε) genotype groups, we observed significant association between IC and statin use (**OR:1.70;1.39-2.09, p = 4.9e-7**) (**Figure 2**).

**Figure 2:**
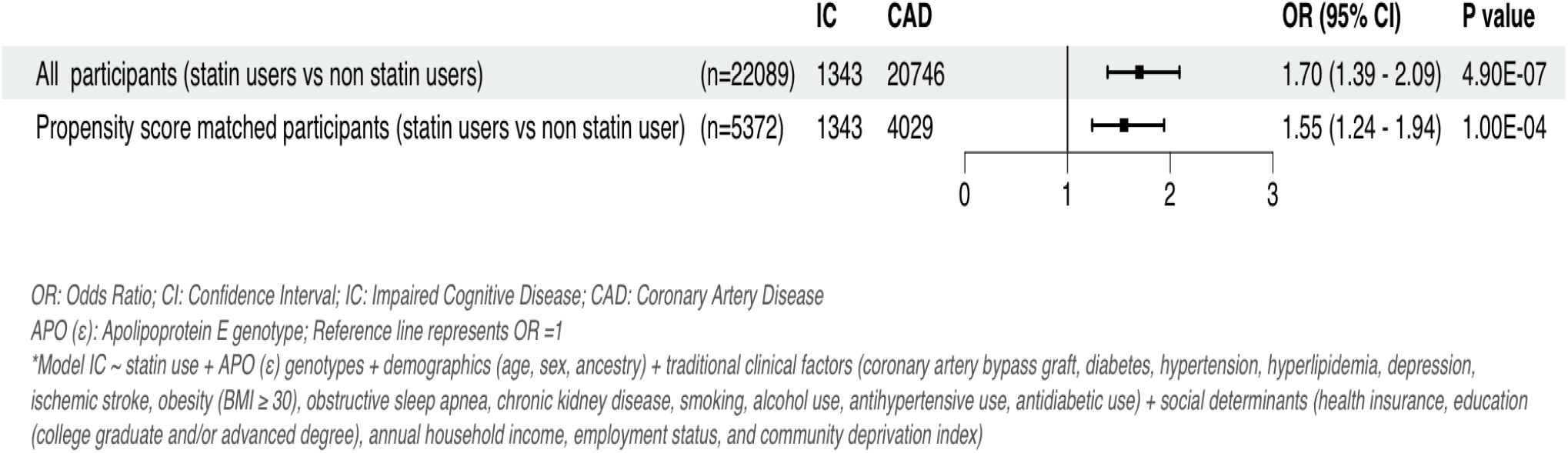
Association between IC and statin use in CAD.

In addition, when we stratified CAD participants based on APO (ε) genotype groups, we observed the highest association between IC and statin use in the APO ε3ε3 group (**OR:2.04;1.53-2.75, p = 1.8e-6**) followed by APO ε3ε4 group (OR:1.61;1.10-2.42, p = 0.01) (**Figure 3**). When we stratified by sex, we observed a stronger magnitude of association between IC and statin use in females (**OR:2.20;1.60-3.06, p=2e-6**) than in males (**1.43;1.10-1.90, p=0.01**) (**Figure 3**).

**Figure 3:**
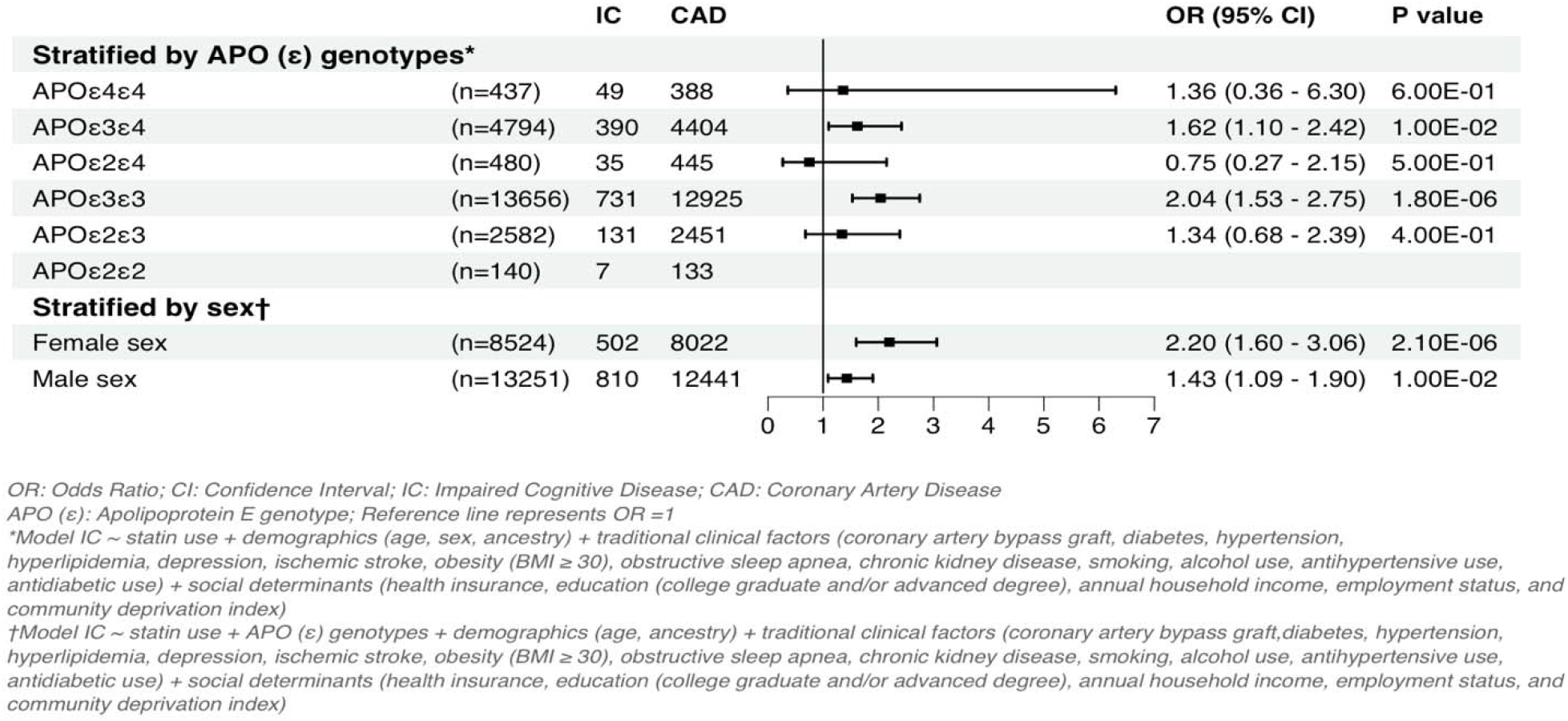
Association between IC and statin use in CAD stratified by APO (ε) genotypes and sex.

### 3) Association of statin use and lipid levels with IC in CAD participants with lipid level data (n=17,050)

Given that lipid levels vary with statin use, we attempted to dissect the association between statin use and IC using baseline, latest, and delta lipid levels (TC, HDL, and LDL) in CAD participants. The mean age of participants when their baseline lipid levels were recorded was lower than their latest lipid levels (62 ± 9.6 yrs vs 73 ± 7.6 yrs, p<0.001).

#### a) Baseline and latest lipid levels

Among CAD participants with lipid levels measured (n=17,050), 75% had baseline lipids assessed before the onset of CAD, and 95% had the latest lipids after the onset of CAD. Among IC (n=1,141), 98% had baseline lipids before the onset of IC, and 70% had the latest lipids after the onset of IC. While the baseline TC (182.7 ± 45.0 mg/dl vs 173.7 ± 43.1 mg/dl, p <0.001) and LDL levels (105.9 ± 39.6 mg/dl vs 96.9 ± 35.2 mg/dl, p<0.001) were higher among statin users compared to non-statin users, and the latest TC (148.3 ± 39.4 mg/dl vs 161.3 ± 41.3 mg/dl, p<0.001) and LDL levels (75.8 ± 31.3 mg/dl vs 86.5 ± 32.8 mg/dl, p<0.001) were lower in statin users compared to non-statin users (**Table 2, Supplement 1**). However, the baseline HDL (48.4 ± 14. 8 mg/dl vs 53.1 ± 16.5 mg/dl, p <0.001) and the latest HDL levels (49.9 ± 15.0 mg/dl vs 53.4 ± 16.3 mg/dl, p<0.001) were lower in statin users as compared to non-statin users (**Table 2, Supplement 1**). We further noted similar trend in both males and females (**Table 3, Table 4, Supplement 1**)

Further, we observed a significant trend for higher proportions of IC among statin users with lower TC, HDL, and LDL compared to non-statin users with higher TC, HDL, and LDL levels (**Table 5, Supplement 1**). When we compared the baseline lipids and latest lipids stratified by statin users across APO (ε) genotype groups, statin users had higher changes in lipids levels across all APO (ε) genotype as compared to non-statin users (**Table 6, Supplement 1**), however, statin users having APO ε3ε3 genotype had the most significant decrease in TC, LDL levels, and increase in HDL levels. (**Table 7, Supplement 1**). When we evaluated the joint association of statin use based on baseline TC, HDL, and LDL levels, we observed consistent association between IC and statin use regardless of the baseline lipid and latest levels as compared to non-statin users (**eFigure 1 eFigure 2, Supplement 1)**.

#### b) Delta lipid levels

The magnitude of lipid level changes from baseline to latest lipids i.e delta lipids were higher statin users as compared non-statin users with higher drops in TC (detalTC: 34 ± 49 mg/dl vs 13 ± 40 mg/dl, p<0.001), LDL levels (deltaLDL: 30 ± 42 mg/dl vs 10 ± 32 mg/dl, p<0.001), and an increase in HDL levels (deltaHDL:2 ± 12 mg/dl. vs 0 ± 11 mg/dl, p<0.001) (**Table 2, Supplement 1**). In our cohort, statin users with the highest lipid variation had the APO ε3ε3 genotype, followed by the APO ε3ε4 genotype (**Table 6, Supplement 1**). When we compared the delta lipids across APO (ε) genotype groups, we observed the most significant statin-associated decrease in TC and LDL levels and the most significant statin-associated increase in HDL levels among the APO ε3ε3 genotype (**Table 7, Supplement 1**).

In multivariable logistic regression analysis, we observed a joint association between statin use and delta lipid levels with IC, regardless of the intensity of decrease in TC and LDL levels, compared with non-statin users (**Figure 4**). However, we observed an increased magnitude of association between IC and statin users having higher HDL increase (**statin: deltaHDL > 10 mg/dl: OR:1.95;1.44-2.66, p=1e-5**) as compared to statin users with lesser HDL increase (**statin: deltaHDL ≤ 10mg/dl: OR:1.61;1.22-2.15, p=8e-4**) (**Figure 4**). The p-value for interaction was 0.005.

**Figure 4:**
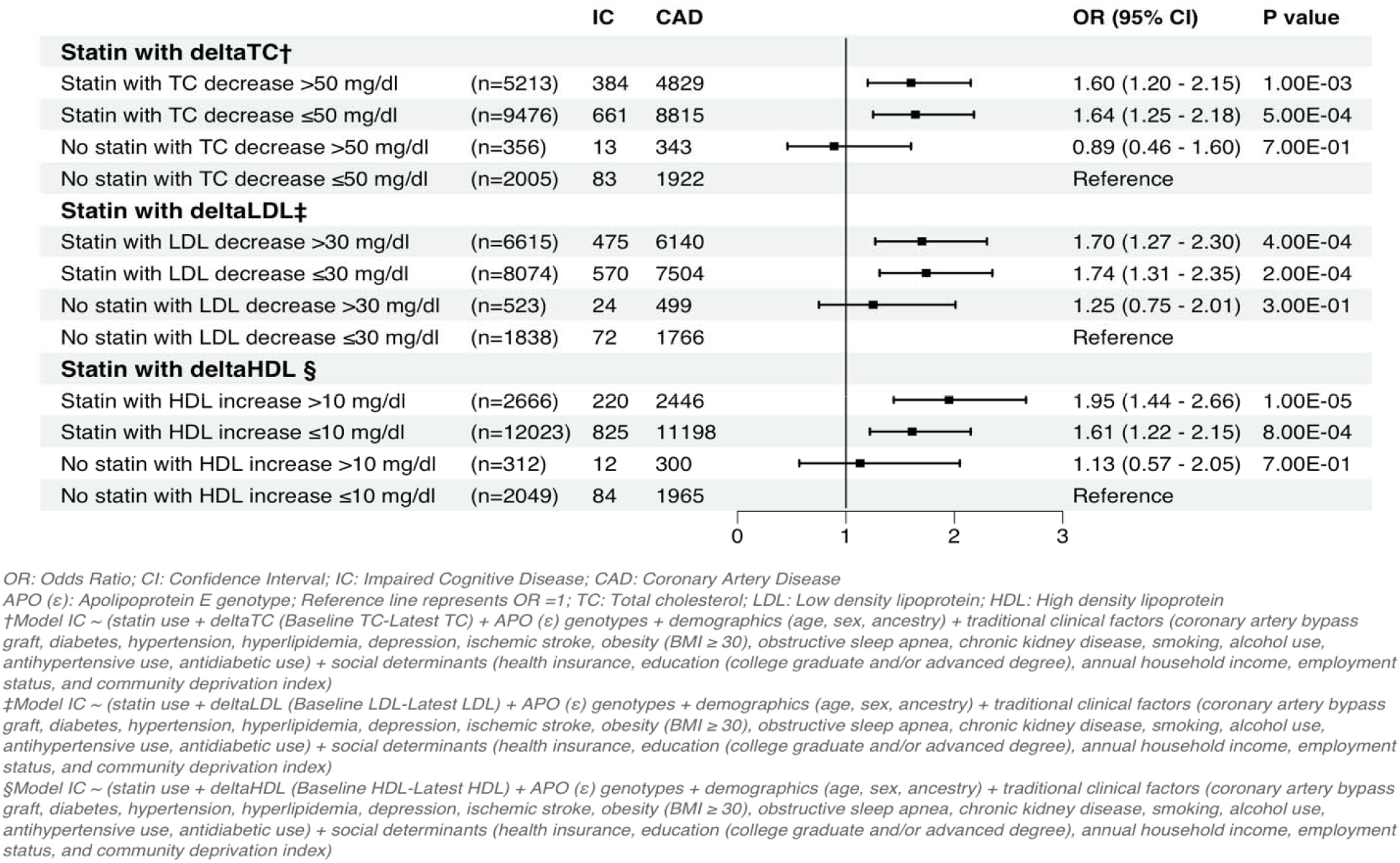
Association between IC and statin use in CAD stratified by changes in lipid levels.

#### c) Sensitivity Analysis

##### 1) Propensity Score Matched Analyses

**e Figure 3 (Supplement 1)** reports the standardized mean difference (SMD) propensity matched plots after matching IC with CAD in all participants and among statin users respectively. The total SMD between IC (n=1,343) and CAD (n=20,746) reduced from 0.67 to 0.05 after matching all variables (IC=1,343, CAD=4,029) (**Table 8, Supplement 1**). After propensity-score matching for demographics, clinical factors, social determinants, and APO (ε) genotypes between IC and CAD, we observed a persistent association between IC and statin use (**OR:1.55;1.24-1.94, p =1e-4**) (**Figure 1**).

##### 2) Association of duration of statin use with IC in statin users

Among CAD participants with statin use, the proportion of IC was higher in statin users with longer duration of statin use (> 8 years vs ≤ 8 years) as compared to shorter duration (8% vs 5%, p<0.001) (**eFigure 4, Supplement 1**). When further noted a signification association between IC and duration of statin use (**OR:1.22;1.06-1.42, p=4e-3**) after adjusting for demographics, clinical factors, social determinants, and APO (ε) genotype groups (**eFigure 4, Supplement 1**). The total SMD between IC (n=1,172) and CAD (n=16,019) reduced from 0.67 to 0.07 after matching all variables (IC=1,172, CAD=3,516) (**eFigure 5 in Supplement 1**). After propensity-score matching, we observed a persistent association between IC and duration of statin use among statin users (**OR:1.21;1.04-1.40, p=0.01**) (**eFigure 4, Supplement 1**).

##### 3) Time to event analysis

In the cox proportional hazard analysis after adjusting for clinical factors, social determinants and APO (ε) genotypes, we observed a higher adjusted HR (1.53;1.20-1.92, p=0.005) for IC with statin use.

##### 4) EtM survey data

We identified (n=812) statin user participants who had completed the optional EtM survey in 2023. When we compared GradCPT (d-prime) scores in CAD participants by duration of statin use, we observed lower scores in those with longer statin use (**Table 9, Supplement 1**). When we performed linear regression to assess the association of duration of statin use with GradCPT scores after adjusting for age, sex, educational status, CDI, and APO (ε) genotype, we observed a consistent negative association of duration of statin use with GradCPT scores (**beta estimate = -0.10, p=0.04**).

## DISCUSSION

In our large retrospective study of diverse AoU CAD participants, we observed an association between IC and statin use, after adjusting for demographics, clinical factors, social determinants, and APO (ε) genotype. In our stratified analysis, we observed the highest magnitude of association between IC and statin use in the APO ε3ε3 group compared to other APO (ε) genotype groups, and in females compared to males. While we did not observe any change in association between IC and statin use, based on the magnitude of decrease in TC and LDL levels, we observed a higher magnitude of association between IC and statin use, with a greater increase in HDL levels. To our knowledge, this is the first study evaluating between IC and statin use based on APO (ε) genotype and serial lipid level evaluations in geriatric CAD.

### Association of statin with IC

The association between statin and IC has been controversial, with mixed results.^6–12,40,41^ While no randomized trial has demonstrated the benefit of statins in reducing cognitive decline, several post hoc trial analyses including PROSPER, ASPREE, TIPS-3 with an average follow up of 4-5 years, have reported no association of statin use with short term cognitive decline.^6,7,12,41^ While all these studies were conducted in patients in the geriatric age group, many of them recruited participants with low baseline vascular risk factor profiles.^7,12^ In the TIPS-3 study, the investigators found no benefit of polypill (antihypertensive + statin) in reducing cognitive decline despite lipid level reduction during the mean follow up of 5 years with one dementia event noted in the polypill group (1/516) and non in placebo group (0/516).^12^ The low baseline vascular risk profile and relatively shorter follow up time in these studies is possibly resulting in low event rates and hence a null association.^7,12^ In comparison, our AoU participants had a mean EHR data length 15 years. More recently, in the UK biobank participants (n=371,019, age >40 yrs) followed over 15 years, statin use was associated with higher risk of AD (HR: 1.19;1.08-1.30) as compared to non-statin users after adjusting for clinical factors, demographics, social determinants, lipid levels, inflammatory biomarkers, and APO (ε) genotype.^8^ In contrast, our AoU cohort focused exclusively on CAD participants and very few large studies have investigated the association of IC and statin use in CAD. The results from the EtM survey data demonstrate an association between the duration of statin use and poor GradCPT scores and our results are consistent with Ye at al., which reported association of statin use with poor neurocognitive test scores in the UK Biobank participants.^8^

### Association of statins with IC based on individual genetic architecture and sex

Consistent with prior CAD cohorts, we observed lower proportions of Alzheimer’s dementia in our CAD participants; however, we may be under-reporting the IC proportions in general, as our EHR follow-up ended in 2023, and due to diagnostic accuracy limitations.^23,28^ Further, our distribution of APO(ε) genotypes is similar with prior reported cohorts.^8,20,23,42^ However, when we stratified our participants based on APO(ε) genotypes, we observed strong association of statin use with IC in the APO ε3ε3 genotype, suggestive of a role of non-APO ε4 carriers in the association of statin use with IC in CAD. Our results somewhat reflect the UK Biobank study, which reported an association of statin use with AD in non-APO ε4 carriers (1.29;1.11-1.49, p<0.001) in the general population.^8^ More recently, drug target Mendelian randomization studies have observed SNVs in statin use related genes such as HMG CoA reductase inhibition, NECTIN-2, APO (ε), PCNX1, DOCK7, and ANGPTL3 to be associated with dementia or poorer neurocognitive test scores.^24,40,43^ While we observed an association of IC with statin use in both males and females, the magnitude of association was stronger in females (**Figure 2**). Our results are again consistent with the UK Biobank study, which reported a higher HR for AD in female statin users (1.37;1.18-1.51).^8^ However, future studies evaluating the association of statin use with IC across sex in CAD based on other comorbid conditions are warranted.

### Interaction of statin use and lipid levels with IC

In our cohort, we observed a steeper decrease in TC and LDL levels and an increase in HDL levels in statin users as compared to non-statin users (**Table 2, Supplement 1**). Our observations are similar to those of Asiimwe et al., who reported serial lipid level changes in the UK Biobank and AoU across statin users.^42^ We observed a strong trend in proportions of IC in statin users with lower latest lipid levels in univariate analysis (**Table 5, Supplement 1**). However, in joint association analysis, IC was associated with statin use regardless of baselines or latest lipid levels. Our results reflect the observations of Chiu et al., who reported that both higher and lower baseline lipid levels (TC and LDL) and steeper lipid variability were associated with the risk of dementia in a large cohort (n=2452, age ≥ 60) in Taiwan.^18^ This pattern was further noted in an independent analysis of ASPREE trial patients by Zhou et al.^19^ In our cohort, we also observed the highest variation in lipid levels in statin users having the APO ε3ε3 genotype, followed by APO ε3ε4 genotype (**Table 7, Supplement 1**) and in our secondary analysis we observed the strongest association between IC and statin use in the APO ε3/ε3 genotype followed by APO ε3/ε4 genotype (**Figure 3**). We further observed an interaction between statin use and delta HDL levels, with a higher magnitude of association between IC and statin use with a steeper increase in HDL levels. Similar observations regarding the risk of IC with increasing HDL levels have been reported in independent cohorts.^18,19^ Lipids are essential for a wide range of body functions, and lower lipid levels and rapid fluctuations may reflect dysfunctional homeostasis and thereby increase the risk of dementia.^18,44^

### Biology of statins and lipids in IC

Statins have a pleiotropic effect on a wide range of plasma lipoproteins, bioactive lipids, and inflammatory biomarkers, and are likely influenced by APO(ε) genotypes.^16,42,45^ For instance, rosuvastatin treatment was associated with a range of upregulation and downregulation of bioactive lipids, including significant downregulation of Palmitoyl Ethanolamide (PEA), which correlated strongly with changes in HDL levels.^45^ PEA in an endogenous lipid mediator with neuroprotective activities, and PEA treatment ameliorated postoperative cognitive decline in mice through attenuation of microglial activation.^46^ More recently, using network toxicology, and in vitro experiments, rosuvastatin has been reported to alter microglial functions via JAK-STAT signaling pathways.^47^ Further fluctuations in plasma cholesterol have been associated with neurocognitive decline and white matter changes.^17–19^ Hence, the biological interaction of plasma lipids, statin use, and APO(ε) genotypes with IC remains complex and can be influenced by comorbidities and lifestyle factors.^16,42^

### Limitations

Our retrospective study design is subject to limitations, including information bias, unmeasured confounding, and measurement bias. However, our study design using the AoU dataset having a standardized data collection model that integrates decades of EHR data for each patient, can somewhat mitigate these biases. Given that AoU participant enrollment required informed consent, participants with severe IC phenotypes at the time of consent were likely excluded, and thus, we may be underreporting IC.^23^ We acknowledge that our IC definition exhibits heterogeneity; however, IC clinical phenotypes often overlap, share heterogeneous neuropathologies, and frequently have a high burden of vascular brain injury.^48^ Further, we increased the likelihood of an accurate diagnosis by restricting to patients aged > 60 years.^23,29^ Similarly, we aimed to capture the entire spectrum of CAD, and our CAD definition shows some heterogeneity. However, our definition is consistent with previously published CAD studies.^22,32^ We used an EHR-based index date of diagnosis for our phenotypes and acknowledge that there could be some discordance between the “true onset of disease” and the EHR-recorded diagnosis date.^49^ We also acknowledge that both CAD and IC are complex phenotypes with significant preclinical evolution that may not be captured adequately in EHR based studies.^23^ While we had adequate power (>80%) for our primary analyses, we did not have enough power for all subgroup analyses. We also acknowledge that there may be heterogeneity in lipid level accuracy due to differences in inter-facility lab protocols, however, EHR-based lipid level measurements from large biobanks like AoU or UK Biobank have been used as predictors in other studies.^8,42^ We also acknowledge that in our primary analysis, we did not account for the type of statin use, and the compliance of statin use.

In conclusion, we observed IC to be significantly associated with statin use, after adjusting for APO (ε) genotype in our AoU CAD participants with age ≥ 60 yrs. In our stratified analysis, we observed the highest magnitude of association between IC and statin use in the APO ε3ε3 group compared to other APO (ε) genotype groups, and in females compared to males.

## Data Availability

All data produced in the present study are available upon reasonable request to the authors

## Code book

Code book retrieving lipid panel, and SBP data will be available on request.

## Acknowledgments

We gratefully acknowledge AllofUs participants for their contributions, without whom this research would not have been possible. We also thank the NIH AllofUs Research Program for making available the participant data examined in this study.

## Author Contributions

The team had full access to all of the data in the study and take responsibility for the integrity of the data and the accuracy of the data analysis.

Concept and Design: Dr.Hariharan

Acquisition, analysis, or interpretation of data: Dr.Hariharan

Drafting of the initial manuscript: Dr.Hariharan

Statistical analysis: Dr.Hariharan, and Dr.Bagheri

Critical review of the manuscript for important intellectual content: Dr.Josee Dupuis, and Dr.Frank Sellke.

Disclaimer: The views expressed are those of the authors and not necessarily those of the funders, NIH, or AllofUS research program.

## Consent Statement

All patients were enrolled in the AoU research program after informed consent.

## Ethics Statement

### Patient and Public Involvement statements

While our outcomes and analysis does not involve direct patient input, our research priorities are in line with the AllofUS precision medicine initiative.

### Ethics approval

Brown University Research Agreements and Contracting Committee reviewed our retrospective analysis proposal and approved the use of AoU researcher workbench. In addition, we have reviewed, completed AoU data access requirements and are in compliance with the data user code of conduct for registered and controlled tier data. There is no human/animal involved in this retrospective de-identified data analysis and hence it does not need to be reviewed and approved by the AoU IRB.

