## Supplement 1 for "Association of cognitive impairment with statin use in coronary artery disease across APO (ε) genotypes in AllofUS": Supplement1_FINAL.docx

Supplement 1 (Index)

1) Case definitions

2) Traditional risks and social determinants definitions.

3) Statistical analysis of CAD cohort with lipid levels and SBP

4)Table 1: Proportions of IC in CAD based on APO (ε) genotype and sex, AllofUS v8, Age ≥60

5) Table 2: Proportions of IC in CAD based on statin use in AllofUS v8, age ≥60

5) Table 2: Lipid level variations based on statin use, AllofUS v8, age ≥60

6) Table 3: Lipid level variations based on statin use in females, AllofUS v8, age ≥60

7) Table 4: Lipid level variations based on statin use in males, AllofUS v8, age ≥60

8) Table 5: Proportions of IC based on statin use and baseline/latest lipid levels, AllofUS v8, age ≥60

9) Table 6: Baseline and latest lipid levels based on statin use, and APO (ε) genotype, AllofUS v8, Age ≥60

10) Table 7: Magnitude of lipid level changes based on statin use, and APO (ε) genotype, AllofUS v8, Age ≥60

9) eFigure 1: Association of statin use with IC in CAD stratified by baseline lipid levels

10) eFigure 2: Association of statin use with IC in CAD stratified by latest lipid levels

11) eFigure 3: Propensity matched plots in all CAD participants (IC with CAD & CAD without IC)

12) Table 6: Baseline characteristics before and after propensity score matching (AllofUS, v8, Age ≥60)

13) eFigure 4: Association of duration of statin use with IC in CAD

14) eFigure 5: Propensity matched plots in all CAD participants with statin use (IC with CAD & CAD without IC)

15) Table 7: Exploring the Mind survey in statin users with CAD

Case definitions

(OMOP Standard Concept Names)

A) Impaired Cognitive disease (IC) had following electronic health records (EHR) diagnoses:

**ICD 9 codes:**

Dementia or MCI: ‘290.0', '290.10', '290.11', '290.12', '290.13', '290.20', '290.21', '290.3', '290.40', '290.41', '290.42', '290.43', ‘290.8’, ‘290.9’, ‘291.2’, ‘294.0’,'294.1', '294.10', '294.11', ‘294.20’, ‘294.21’,'331.0', ‘331.7’,'331.11', '331.19',’331.2’,’331.83’

**ICD 10 codes:**

Dementia or MCI: 'F01.50', 'F01.51', ‘F01.52’, ‘F01.A0’, ‘F01.B0’, ‘F01.C4’,‘F02.80’, ‘F02.81’,’F02.811’, ‘F02.818’, ‘F02.A0’, ‘F02.A18’, ‘F02.B0’, ‘F02.B3’, ‘F02.B4’, ‘F03.90', 'F03.91', ‘F03.911’, ‘F03.918’, ‘F03.92’, ‘F03.92’, ‘F03.93’, ‘F03.94’, ‘F03.A0’, ‘F03.A18’, ‘F03.B18’, ‘F03.C3’, ‘G30.1’, ‘G30.8’, 'G30.9', 'G31.01', 'G31.84', ‘G31.84’, 'F10.27'

- Alzheimer's disease
- Amnestic disorder
- Dementia
- Dementia associated with alcoholism
- Dementia associated with another disease
- Dementia with behavioral disturbance
- Frontotemporal dementia
- Mild cognitive disorder
- Mild dementia
- Minimal cognitive impairment
- Moderate dementia
- Multi-infarct dementia with depression
- Multi-infarct dementia, uncomplicated
- Presenile dementia with depression
- Primary degenerative dementia of the Alzheimer type, presenile onset
- Primary degenerative dementia of the Alzheimer type, senile onset
- Senile degeneration of brain
- Senile dementia with depression
- Uncomplicated presenile dementia
- Uncomplicated senile dementia
- Vascular dementia with behavioral disturbance
- Vascular dementia without behavioral disturbance

B) Coronary Artery Disease (CAD) had following electronic health records (EHR) diagnoses:

**ICD 9 codes:** ‘413.0’, ‘413.1’, ‘413.9’, ‘414’,’414.0’, ‘414.00’, ‘414.01’, ‘414.02’, ‘414.03’, ‘414.04’

‘414.05’, ‘414.07’, ‘414.10’, ‘414.11’, ‘414.12’, ‘414.19’, ‘414.2’, ‘414.3’, ‘414.4’, ‘414.8’, ‘414.9’

**ICD 10 codes:** ‘I20’, ‘I20.0’, ‘I20.1’, ‘I20.8’, ‘I20.9’, ‘I21’, ‘I21.01’, ‘I21.02’, ‘I21.09’

‘I22.1’, ‘I22.2’, ‘I23.1’, ‘I23.3’, ‘I23.5’, ‘I23.6’, ‘I23.8’, ‘I24.0’, ‘I24.1’, ‘I24.8’,

‘I24.9’, ‘I25’, ‘I25.1’, ‘I25.10’, ‘I25.11’, ‘I25.110’, ‘I25.111’, ‘I25.118’ , ‘I25.119’, ‘I25.2’

‘I25.3’, ‘I25.41’, ‘I25.42’, ‘I25.5’, ‘I25.6’, ‘I25.700’, ‘I25.701’, ‘I25.708’, ‘I25.709’, ‘I25.710’,

‘I25.718’, ‘I25.719’, ‘I25.720’, ‘I25.721’, ‘I25.729’, ‘I25.730’, ‘I25.738’, ‘I25.758’, ‘I25.759’, ‘I25.760’

‘I25.768, ‘I25.790’, ‘I25.810’, ‘I25.812’, ‘I25.82’, ‘I25.83’, ‘I25.84’, ‘I25.89’, ‘I25.9’

1) Chronic total occlusions of coronary artery
2) Acute ischemic heart disease
3) Chronic ischemic heart disease
4) Coronary artery thrombosis
5) Coronary arteriosclerosis
6) Acute coronary artery occlusion not resulting in myocardial infarction
7) Coronary artery atherosclerosis
8) Coronary artery bypass graft finding
9) Coronary thrombosis not resulting in myocardial infarction
10) Myocardial infarction due to demand ischemia
11) Angina pectoris
12) Acute ST elevation myocardial infarction
13) Coronary artery bypass graft occlusion
14) Atherosclerosis of coronary artery without angina

Definitions of predictors and co-variates

(OMOP Standard Concept Names with SNOMED Concept ID)

A) Statin drugs:

Concept ID: 21601855

B) Lipid level data:

1) Total Cholesterol (TC):

Concept ID: 4260765, 44791053, 4190897, 40484105, 4008265,3027114,40757569,2212267,4260765, 44791053, 4190897, 40484105, 4008265, 3037598, 40757569, 4260765, 44791053, 4190897, 3002651, 2212267,44787078, 40484105, 4008265, 40780291,3027114, 40757569,2212267, 3024723,3033364, 3024723, 3033364, 3015232

2) Low density lipoprotein (LDL):

Concept ID: 3008631,3028437,3009966, 3035899, 3028288,3053341, 2212450, 3049222, 3050458, 40760809, 3035009, 3049237, 3052982, 3028288, 3046493,3001308, 3045323, 3009966, 4191837, 4041721, 4042062, 40795800, 3033200, 42870529, 3050730, 3030437, 3038988, 3008631, 3035899, 4210878, 4041556, 4042061, 4042758

3) Hight density lipoprotein (HDL):

Concept ID: 3007070,3050630,3032633,3011163, 3009718, 4041557, 3053286, 4041720, 4042059, 4042081, 4055665, 4076704, 4101713, 4195503, 3003767, 3005561, 3033011, 3007070, 3009718,3 011884, 3013473, 3015204, 3020107, 3020189, 3022449,3023574,3023602,3023752,3024401,3030792, 40761042, 40757503, 40782589,3032771, 3033190, 3033638, 3034482,3040324,3040815, 40759254, 3050988,40798534, 40795258, 40795251, 40785417, 40789378, 40795256, 40782086, 40788729, 3033190, 3020189, 3023602, 3009718, 40761043, 40759253, 42868674, 40758736, 3047111,3050038, 40757599, 4021289, 40778721, 40775398, 40795257, 40795254, 40788728, 40795253, 40785416, 40782761, 40778719

C) Systolic Blood Pressure (SBP):

Concept ID: 4232915, 4292062, 4161413, 4248525, 4197167, 37394652, 37396683, 3012311, 3025407, 3033616, 3025048, 3025957, 3011870, 3035856, 21492239, 3018586, 903114, 903118, 46284421, 4152194, 3013880, 3009395, 3011579, 3010693, 3006833, 3013443, 903130, 903109,3004249

D) Traditional clinical factors:

1) Body Mass Index (BMI):

- Earliest BMI value obtained from electronic medical records (3038553)

2) Depression: (440383, 4152280)

- Acute depression
- Atypical depressive disorder
- Bipolar affective disorder, current episode depression
- Chronic depression
- Chronic depressive personality disorder
- Depressive disorder in remission
- Dysthymia
- Major depressive disorder
- Mild, moderate, severe and recurrent depression
- Schizoaffective disorder, depressive type

3) Ischemic stroke: (4111710, 443454, 381316, 4153352, 4310996, 4046360)

- Brainstem stroke syndrome
- Cerebral infarction
- Cerebrovascular accident
- Embolic stroke
- Ischemic stroke
- Lacunar infarction

4) Hypertension: (312648, 4028741, 4167358, 320128, 317898, 4209293)

- Benign hypertension
- Essential hypertension
- Malignant essential hypertension
- Diastolic and Systolic hypertension

5) Hyperlipidemia: (44834563, 432867, 35207065, 438720, 35207062, 35207061)

- Endogenous hyperlipidemia
- Hypercholesterolemia
- Hypertriglyceridemia
- Mixed hyperlipidemia
- Secondary hyperlipidemia

6) Diabetes: (201820, 44833365, 1567940, 201826, 1567956)

- Diabetes during pregnancy
- Diabetes without complication
- Secondary diabetes
- Type 1 and 2 diabetes

9) Alcohol use: (4218106)

- Alcohol abuse
- Alcohol dependence
- Chronic alcoholism in remission
- Chronic continuous alcoholism
- Episodic chronic alcoholism

10) Smoking: (4209423)

- Nicotine dependence
- Nicotine dependence in remission
- Tobacco dependence syndrome

11) Coronary artery bypass graft (CABG):

a) Aortocoronary bypass graft present (42537729)

- Coronary artery bypass graft finding (312922)
- Coronary artery bypass graft present (42537730)

b) Arteriosclerosis of coronary artery bypass graft (443563)

12) Obstructive sleep apnea: (442588)

- Mixed sleep apnea
- Obstructive sleep apnea of adult

13) Chronic kidney disease: (46271022, 443614, 443601, 443597, 45763854, 443611, 443612, 4030520)

- Chronic kidney disease stage 1-5
- End-stage renal failure on dialysis.

12) Antihypertensive drugs: (21600381)

- Betablockers (21601664)
- Renin-angiotensin system inhibitors (21601782)
- Diuretics (21601461)
- Peripheral vasodilators (21601560)
- Calcium channel blockers (21601744)

13) Antidiabetic drugs

- Fast acting insulin (51428, 400008, 86009, 314686, 253182, 221109)
- Intermediate acting insulin (314684)
- Long acting insulin (1670007, 139825, 274783)
- Insulin analogues for inhalation (631657)
- Biguanides (6809, 8129)
- Sulfonylurea (2404, 25789, 4821, 4815, 10633, 10635)
- Alphaglucosidase inhibitors (16681, 30009)
- Thiazolidiendiones (33738, 84108, 72610)
- Dipeptidyl peptidase 4 inhibitors (1368001, 1100699, 857974, 593411)
- Glucagon like peptide-1 inhibitors (1534763, 1551291, 60548, 475968, 1440051, 1991302)
- Sodium-glucose- cotransport 2 inhibitors (1373458, 1488564, 1545653, 1992672)

C) Social determinants:

1) Annual household income: (1585375)

- Skipped/prefer not to answer (Group = 0)
- Annual income (USD≤ $25,000, Group = 1)
- Annual income (USD> $25,000 and ≤ $75,000, Group = 2)
- Annual income (USD> $75,000,Group = 3)

2) Employment: (1585952)

- Skipped/prefer not to answer (Group = 0)
- Unable to work or out of work (Group = 1)
- Retired (Group = 2)
- Employed (Group = 3)

3) Health insurance status (585386):

- No/Skip/Prefer not to answer (Group = 0)
- Yes (Group = 1)
- Don’t know (Group = 2)

4) Community Deprivation index (composite score based on the following census tract variables):

- Poverty fraction: Fraction of households with income below poverty level within the past 12 months
- Median income: Median household income in the past 12 months in 2017 inflation-adjusted dollars
- Fraction high school: Fraction of population 25 and older with educational attainment of at least high school graduation (includes GED equivalency)
- Fraction insured: fraction of population with health insurance
- Fraction SNAP: Fraction of households receiving public assistance income or food stamps/SNAP in the past 12 months
- Fraction vacant: Fraction of houses that are vacant

The CDI ranges from 0-1, with higher values indicating greater deprivation, and we subsequently categorized CDI into low ( ≤ 0.29), medium (0.29-0.34), and high tertiles (≥ 0.35) for the purpose of our analysis

CAD cohort with lipid levels and SBP

Step 1: We curated lists of AoU v8 participants with recorded lipid levels in the EHR, based on concept codes corresponding to TC, HDL, and LDL (Details in Supplement 1). Subsequently, we ordered (ascending and descending) individual lipid data by reported EHR dates to extract the oldest lipid levels (baseline TC, HDL, and LDL) and the most recent lipid levels (latest TC, HDL, and LDL). We merged the AoU participant data with baseline and recent lipid data into a joint lipid data set (n=143,540). While merging, we ensured that all baseline lipids and all recent lipids had the same reported dates. To add SBP data, we curated a list of AoU participants with SBP recorded at enrollment (participant-provided information) based on concept ID (Details in Supplement 1). We obtained the lowest SBP value for each participant across multiple measurements. We merged the SBP data and joint lipid data set (n=143,540) with our CAD participant list (n=22,039) to include unique CAD participants with lipids and SBP data (n=17,050) (**Figure 1**). We subsequently performed univariate analysis comparing baseline and recent lipids across statin users and non-statin users, in all participants and in each APO (ε) genotype group to assess the trend of lipid level changes based on APO (ε) genotype.

Step 2: Given the lipid level targets in CAD vary based on individual risks, and extreme variation in lipid levels are associated with IC, we categorized baseline and latest lipids as TC130 (≥ 130mg/dl or < 130 mg/dl), HDL40 (≥ 40mg/dl or < 40mg/dl) and LDL (≥55 mg/dl, or <55 mg/dl) to better understand the joint association of these dichotomous lipid categories and statin use with IC in CAD.^5,10,14,15,18^ We subsequently performed univariate analysis comparing proportions of IC across statin users stratified by TC130, HDL40, and LDL55, respectively (baseline and latest). In order to assess the joint effect of statin use with individual lipid level targets (baseline and latest), we introduced joint effect terms of statin use with TC130 (statin: TC130), HDL40 (statin: HDL40), and LDL55 (statin: LDL55) and ran similar logistic regression models in all CAD participants.

Step 3: Since fluctuations in individual lipid levels have been associated with IC and can be influenced by statin therapy, we wanted to assess the association between statin use and IC based on the magnitude of the difference between baseline and latest lipid levels.^18,19^ For this analysis, we quantified the magnitude of difference between baseline lipids (first TC, HDL, and LDL) and latest lipids (latest TC, HDL, and LDL) as delta TC, delta HDL, and delta LDL, respectively. We subsequently categorized delta lipids as deltaTC50 (> 50 mg/dl, reduction), deltaHDL10 (>10 mg/dl, increase), and deltaLDL30 (>30 mg/dl, reduction). We assessed the association between IC and statin use based on delta lipids using joint effect terms (statin: deltaTC50, statin: HDL10, and statin: deltaLDL30), and performed a similar logistic regression analysis in CAD participants after adjusting for demographics, traditional risk factors, social determinants, and APO (ε) genotypes.

For the univariate and joint association analysis of lipid level categories and statin use, we defined Bonferroni-adjusted significance at 0.004 (0.05/12) based on 4 subcategory association testing for each joint effect term in TC, HDL, and LDL regression models, respectively.

Sensitivity Analysis

a) Propensity-score matched analyses:

Given that the traditional clinical factors in IC can confound the association of between IC and statin use and CAD, we propensity-matched the IC (with CAD) to CAD (without IC) for demographics, traditional risk factors, social determinants, and APO (ε) genotypes. We used nearest-neighborhood matching, maintaining a case (IC with CAD) and treated-control (CAD without IC) ratio of 1:3.^40^ We repeated our primary logistic regression models in the matched group to assess the association of between IC and statin use after adjusting for demographics, traditional risk factors, social determinants, and APO (ε) genotypes. We defined significance at p < 0.05 for this analysis.

b) Duration of statin use:

To assess the association between IC and duration of statin use in CAD, we calculated the total years of statin use for each CAD participant as the difference between the index year of statin therapy recorded in the EHR and 2023 (when the EHR data were released). Subsequently, we categorized each CAD participant as having used statins for >8 years or ≤8 years. We used 8 years based on the mean duration of statin use (8.4 years) observed among CAD participants prior to their first observed IC event. Subsequently, we assessed the association between IC and duration of statin use (> 8 years vs ≤ 8 years) by running similar logistic regression models in all CAD participants while adjusting for demographics, traditional risk, social determinants, and APO (ε) genotypes. We repeated our primary logistic regression models after propensity score matching to assess the association between IC and duration of statin use (> 8 years vs ≤ 8 years) and IC. We defined significance at p < 0.05 for this analysis.

c) EtM survey data analysis:

Given the heterogeneity of EHR-based IC definitions, we intended to assess the association between statin use duration and neurocognitive test scores. For this analysis, we restricted to statin users who completed this optional EtM survey in 2023. Subsequently, we compared GradCPT scores in CAD participants with different durations of statin use (>8 years vs ≤ 8 years) using a t-test. Finally, we assessed the association between GradCPT scores and duration of statin use using linear regression while adjusting for age, sex, educational status, CDI, and APO (ε) genotype. We defined significance at p < 0.05.

| Table 1: Proportions of IC in CAD based on APO (ε) genotype and sex, AllofUS v8, Age ≥60 | | | |
| --- | --- | --- | --- |
| CAD participants (n=22,089) | | | |
|  | No statin  (n=4,898) | Yes statin  (n=17,191) | P Value* |
| IC (n=1348) | 171/4898 (3.5%) | 1172/17191 (6.8%) | **p<0.001** |
| Stratified by APO (ε) genotype | | | |
| APOε2ε2 (n=140) | 2/31 (6.5%) | 5/109 (4.6%) | p=0.67 |
| APOε2ε3 (n=2,582) | 18/629 (2.9%) | 113/1953 (5.8%) | **p=0.004** |
| APOε3ε3 (n=13,656) | 85/3003 (2.8%) | 646/10653 (6.1%) | **p<0.001** |
| APOε2ε4 (n=480) | 12/126 (9.5%) | 23/354 (6.5%) | p=0.26 |
| APOε3ε4 (n=4,794) | 46/1020 (4.5%) | 344/3774 (9.1%) | **p<0.001** |
| APOε4ε4 (n=437) | 8/89 (9.0%) | 41/348 (11.8%) | p=0.46 |
| Stratified by sex | | | |
| Females (n=8,524) | 63/2153 (2.9%) | 439/6371 (6.9%) | **p<0.001** |
| Males (n=13,251) | 105/2682 (3.9%) | 705/10569 (6.7%) | **p<0.001** |
| *Bold values with higher proportions having p value ≤0.05  CAD: Coronary Artery Disease  IC: Impaired cognitive disease defined as all-cause dementia or mild cognitive impairment | | | |

| Table 2: Proportions of IC in CAD based on statin use in AllofUS v8, age ≥60 | | | |
| --- | --- | --- | --- |
| CAD participants (n=22,089) | | | |
|  | No statin  (n=4,898) | Yes statin  (n=17,191) | P Value* |
| IC (n=1348) | 171 (3.5%) | 1172 (6.8%) | **p<0.001** |
| Stratified by Type of IC |  |  |  |
| Mild cognitive impairment | 86/4898 (1.7%) | 629/17191 (3.6%) | **p<0.001** |
| Alzheimer’s dementia | 6/4898 (0.1%) | 52/17191 (0.3%) | **p=0.04** |
| Vascular dementia | 9/4898 (0.2%) | 63/17191 (0.4%) | p=0.06 |
| Other dementia | 70/4898 (1.4%) | 428/17191 (2.5%) | **p<0.001** |
| * Bold values with higher proportions having p value ≤0.05  CAD: Coronary Artery Disease;  IC: Impaired cognitive disease defined as all-cause dementia or mild cognitive impairment | | | |

| Table 2: Lipid level variations based on statin use, AllofUS v8, age ≥60 | | | | | |
| --- | --- | --- | --- | --- | --- |
| CAD participants with lipid levels (n=17,050) | | | | | |
| No statin  (n=2,361) | | P Value | Yes statin  (n=14,689) | | P Value* |
| Participant age at baseline lipids (mean ± SD, years) | Participant age at latest lipids (mean ± SD, years) |  | Participant age at baseline lipids (mean ± SD, years) | Participant age at latest lipids (mean ± SD, years) |  |
| 65.6 (±8.9) years | 72.26 (±7.7) years | **p<0.001** | 61.55 (± 9.6) years | 73.4 (±7.5) years | **p<0.001** |
| Baseline TC (mean ± SD, mg/dl) | Latest TC (mean ± SD, mg/dl) |  | Baseline TC (mean ± SD, mg/dl) | Latest TC (mean ± SD, mg/dl) |  |
| 174 ± 43 mg/dl | 161 ± 41 mg/dl | **p<0.001** | 183 ± 45 mg/dl | 148 ± 39 mg/dl | **p<0.001** |
| Delta TC (TC baseline-TC latest, mean ± SD, mg/dl) | |  | Delta TC (TC baseline-TC latest, mean ± SD, mg/dl) | |  |
| 13 ± 40 mg/dl | | NA | 34 ± 49 mg/dl | | **p<0.001** |
| Baseline HDL (mean ± SD, mg/dl) | Latest HDL (mean ± SD, mg/dl) |  | Baseline HDL (mean ± SD, mg/dl) | Latest HDL (mean ± SD, mg/dl) |  |
| 53 ± 17 mg/dl | 53 ± 16 mg/dl | p=0.41 | 48 ± 15 mg/dl | 50 ± 15 mg/dl | **p<0.001** |
| Delta HDL (HDL baseline-HDL latest, mean ± SD, mg/dl) | |  | Delta HDL (HDL baseline-HDL latest, mean ± SD, mg/dl) | |  |
| 0 ± 11 mg/dl | | NA | -(2) ± 12 mg/dl | | **p<0.001** |
| Baseline LDL (mean ± SD, mg/dl) | Latest LDL (mean ± SD, mg/dl) |  | Baseline LDL (mean ± SD, mg/dl) | Latest LDL (mean ± SD, mg/dl) |  |
| 97 ± 35 mg/dl | 87 ± 33 mg/dl | **p<0.001** | 106 ± 40 mg/dl | 76 ± 31 mg/dl | **p<0.001** |
| Delta LDL (LDL baseline-HDL latest, mean ± SD, mg/dl) | |  | Delta LDL (LDL baseline-LDL latest, mean ± SD, mg/dl) | |  |
| 10± 32 mg/dl | | NA | 30 ± 42 mg/dl | | **p<0.001** |
| * Bold values with higher proportions having p value ≤0.05  CAD: Coronary Artery Disease  IC: Impaired cognitive disease defined as all-cause dementia or mild cognitive impairment  TC: Total cholesterol; HDL: High density lipoprotein; LDL: Low density lipoprotein; SBP: Systolic blood pressure | | | | | |

| Table 3: Lipid level variations based on statin use in females, AllofUS v8, age ≥60 | | | | | |
| --- | --- | --- | --- | --- | --- |
| Female CAD participants with lipid levels (n=6,548) | | | | | |
| No statin  (n=1,146) | | P Value | Yes statin  (n=5,402) | | P Value* |
| Baseline TC (mean ± SD, mg/dl) | Latest TC (mean ± SD, mg/dl) |  | Baseline TC (mean ± SD, mg/dl) | Latest TC (mean ± SD, mg/dl) |  |
| 182 ± 47 mg/dl | 171 ± 37 mg/dl | **p<0.001** | 192 ± 44 mg/dl | 160 ± 40 mg/dl | **p<0.001** |
| Baseline HDL (mean ± SD, mg/dl) | Latest HDL (mean ± SD, mg/dl) |  | Baseline HDL (mean ± SD, mg/dl) | Latest HDL (mean ± SD, mg/dl) |  |
| 59 ± 16 mg/dl | 59 ± 16 mg/dl | p=0.71 | 54 ± 16 mg/dl | 56 ± 16 mg/dl | **p<0.001** |
| Baseline LDL (mean ± SD, mg/dl) | Latest LDL (mean ± SD, mg/dl) |  | Baseline LDL (mean ± SD, mg/dl) | Latest LDL (mean ± SD, mg/dl) |  |
| 100 ± 36 mg/dl | 91 ± 32 mg/dl | **p<0.001** | 111 ± 39 mg/dl | 81 ± 32 mg/dl | **p<0.001** |
| * Bold values with higher proportions having p value ≤0.05  CAD: Coronary Artery Disease  IC: Impaired cognitive disease defined as all-cause dementia or mild cognitive impairment  TC: Total cholesterol; HDL: High density lipoprotein; LDL: Low density lipoprotein; SBP: Systolic blood pressure | | | | | |

| Table 4: Lipid level variations based on statin use in males, AllofUS v8, age ≥60 | | | | | |
| --- | --- | --- | --- | --- | --- |
| Male CAD participants with lipid levels (n=10,255) | | | | | |
| No statin  (n=1,188) | | P Value | Yes statin  (n=9,067) | | P Value* |
| Baseline TC (mean ± SD, mg/dl) | Latest TC (mean ± SD, mg/dl) |  | Baseline TC (mean ± SD, mg/dl) | Latest TC (mean ± SD, mg/dl) |  |
| 160 ± 41 mg/dl | 146 ± 37 mg/dl | **p<0.001** | 176 ± 44 mg/dl | 139 ± 36 mg/dl | **p<0.001** |
| Baseline HDL (mean ± SD, mg/dl) | Latest HDL (mean ± SD, mg/dl) |  | Baseline HDL (mean ± SD, mg/dl) | Latest HDL (mean ± SD, mg/dl) |  |
| 47 ± 15 mg/dl | 47 ± 15 mg/dl | p=0.75 | 44 ± 12 mg/dl | 46 ± 13 mg/dl | **p<0.001** |
| Baseline LDL (mean ± SD, mg/dl) | Latest LDL (mean ± SD, mg/dl) |  | Baseline LDL (mean ± SD, mg/dl) | Latest LDL (mean ± SD, mg/dl) |  |
| 91 ± 34 mg/dl | 77 ± 31 mg/dl | **p<0.001** | 102 ± 39 mg/dl | 71 ± 28 mg/dl | **p<0.001** |
| * Bold values with higher proportions having p value ≤0.05  CAD: Coronary Artery Disease  IC: Impaired cognitive disease defined as all-cause dementia or mild cognitive impairment  TC: Total cholesterol; HDL: High density lipoprotein; LDL: Low density lipoprotein; SBP: Systolic blood pressure | | | | | |

| Table 5: Proportions of IC based on statin use and baseline/latest lipid levels, AllofUS v8, age ≥60 | | | | | |
| --- | --- | --- | --- | --- | --- |
| Proportions of IC | | | | | |
|  | No statin  (n=2,361) | | Yes statin  (n=14,689) | | P Value* |
| IC (n=1,141) | 96 (4%) | | 1,045 (7.1%) | | **p<0.001** |
|  | **Stratified by baseline lipid levels** | | | |  |
|  | **TC** ≥ **130mg/dl** | **TC < 130mg/dl** | **TC** ≥ **130mg/dl** | **TC < 130mg/dl** |  |
| TC | 71/1986  (3.6%) | 25/375  (6.7%) | 913/12930  (7.1%) | 132/1759  (7.5%) | **p<0.001**† |
|  | **HDL** ≥ **40mg/dl** | **HDL < 40mg/dl** | **HDL** ≥ **40mg/dl** | **HDL < 40mg/dl** |  |
| HDL | 73/1881  (3.9%) | 23/480  (4.8%) | 720/10205  (7.1%) | 325/4484  (7.2%) | **p<0.001**† |
|  | **LDL** ≥ **55 mg/dl** | **LDL < 55 mg/dl** | **LDL** ≥ **55mg/dl** | **LDL < 55 mg/dl** |  |
| LDL | 82/2110  (3.9%) | 14/251  (5.6%) | 949/13,469  (7.0%) | 96/1220  (7.9%) | **p<0.001**† |
|  | **Stratified by latest lipid levels** | | | |  |
|  | **TC** ≥ **130mg/dl** | **TC < 130mg/dl** | **TC** ≥ **130mg/dl** | **TC < 130mg/dl** |  |
| TC | 61/1802  (3.4%) | 35/559  (6.3%) | 648/9544  (6.8%) | 397/5145  (7.7%) | **p<0.001**† |
|  | **HDL** ≥ **40mg/dl** | **HDL < 40mg/dl** | **HDL** ≥ **40mg/dl** | **HDL < 40mg/dl** |  |
| HDL | 72/1888  (3.8%) | 24/473  (5.1%) | 763/10845  (7.0%) | 282/3562  (7.3%) | **p<0.001**† |
|  | **LDL** ≥ **55 mg/dl** | **LDL < 55 mg/dl** | **LDL** ≥ **55mg/dl** | **LDL < 55 mg/dl** |  |
| LDL | 76/1970  (3.9%) | 20/371  (5.1%) | 745/10850  (6.9%) | 300/3839  (7.8%) | **p<0.001**† |
| * Bold values with higher proportions having p value ≤0.05  † p for trend test  CAD: Coronary Artery Disease  IC: Impaired cognitive disease defined as all-cause dementia or mild cognitive impairment  TC: Total cholesterol; HDL: High density lipoprotein; LDL: Low density lipoprotein; SBP: Systolic blood pressure | | | | | |

| Table 6: Baseline and latest lipid levels based on statin use, and APO (ε) genotype, AllofUS v8, Age ≥60 | | | | | | | | | | |
| --- | --- | --- | --- | --- | --- | --- | --- | --- | --- | --- |
| CAD participants with lipid levels (n=17,050) | | | | | | | | | | |
| APO (ε) genotype | Statin therapy (n) | Baseline TC (mean ± SD, mg/dl) | Latest TC (mean ± SD, mg/dl) | p value | Baseline HDL (mean ± SD, mg/dl) | Latest HDL (mean ± SD, mg/dl) | p value | Baseline LDL (mean ± SD, mg/dl)) | Latest LDL (mean ± SD, mg/dl) | **p**  **value** |
| APOε2ε2 | No  (n=12) | 170 ± 39 mg/dl | 160 ± 47 mg/dl | 5e-1 | 54 ± 16 mg/dl | 51 ± 13 mg/dl | 6e-1 | 76 ± 32 mg/dl | 63 ± 25 mg/dl | 3e-1 |
| APOε2ε2 | Yes (n=94) | 188 ± 54 mg/dl | 139 ± 42 mg/dl | **7e-11** | 49 ± 13 mg/dl | 50 ± 14 mg/dl | 4e-1 | 90 ± 42 mg/dl | 62 ± 28 mg/dl | **3e-7** |
| APOε2ε3 | No  (n=335) | 169 ± 39 mg/dl | 160 ± 39 mg/dl | **4e-3** | 55 ± 17 mg/dl | 56 ± 18 mg/dl | 2e-1 | 90 ± 30 mg/dl | 81 ± 29 mg/dl | **7e-5** |
| APOε2ε3 | Yes  (n=1697) | 174 ± 44 mg/dl | 141±39 mg/dl | **6e-112** | 49 ± 15 mg/dl | 50 ± 15 mg/dl | **1e-3** | 95 ± 37 mg/dl | 68 ± 29 mg/dl | **1e-119** |
| APOε3ε3 | No  (n=1423) | 174 ± 43 mg/dl | 161 ± 41 mg/dl | **2e-15** | 53 ± 16 mg/dl | 53± 16 mg/dl | 9e-1 | 97 ± 35 mg/dl | 87 ± 33 mg/dl | **3e-16** |
| APOε3ε3 | Yes  (n=9100) | 183 ± 45 mg/dl | 148 ± 39 mg/dl | **0e-0** | 49 ± 15 mg/dl | 50 ± 15 mg/dl | **1e-11** | 106 ± 39 mg/dl | 76 ± 31 mg/dl | **0e-0** |
| APOε2ε4 | No  (n=69) | 169 ± 46 mg/dl | 157 ± 41 mg/dl | 1e-1 | 51 ± 17 mg/dl | 53 ± 16 mg/dl | 3e-1 | 89 ± 34 mg/dl | 79 ± 29 mg/dl | 7e-2 |
| APOε2ε4 | Yes  (n=306) | 179 ± 43 mg/dl | 144 ± 39 mg/dl | **1e-23** | 48 ± 15 mg/dl | 51 ± 16 mg/dl | 6e-2 | 102 ± 39 mg/dl | 72 ± 31 mg/dl | **8e-24** |
| APOε3ε4 | No  (n=479) | 177 ± 47 mg/dl | 162 ± 43 mg/dl | **2e-7** | 52 ± 17 mg/dl | 52 ± 16 mg/dl | 9e-1 | 101 ± 38 mg/dl | 89 ± 35 mg/dl | **2e-6** |
| APOε3ε4 | Yes  (n=3195) | 186 ± 45 mg/dl | 152 ± 41 mg/dl | **1e-212** | 48 ± 15 mg/dl | 49 ± 15 mg/dl | **7e-6** | 110 ± 40 mg/dl | 80 ± 35 mg/dl | **1e-221** |
| APOε4ε4 | No  (n=43) | 184 ± 52 mg/dl | 171 ± 38 mg/dl | 1e-1 | 52 ± 17 mg/dl | 52 ± 16 mg/dl | 9e-1 | 110 ± 41 mg/dl | 99 ± 38 mg/dl | 1e-1 |
| APOε4ε4 | Yes  (n=297) | 193 ± 47 mg/dl | 155 ± 38 mg/dl | **7e-25** | 47 ± 14 mg/dl | 50 ± 16 mg/dl | 7e-2 | 119 ± 45 mg/dl | 83 ± 31 mg/dl | **2e-26** |
| * Bold values having Bonferroni adjusted student t.test p value ≤0.004 (0.05/12)  CAD: Coronary Artery Disease  TC: Total cholesterol; HDL: High density lipoprotein; LDL: Low density lipoprotein | | | | | | | | | | |

| Table 7: Magnitude of lipid level changes based on statin use, and APO (ε) genotype, AllofUS v8, Age ≥60 | | | | | | | |
| --- | --- | --- | --- | --- | --- | --- | --- |
| CAD participants with lipid levels (n=17,050) | | | | | | | |
| APO (ε) genotype | Statin therapy (n) | Delta TC (TC baseline-TC latest, mean ± SD, mg/dl) | p value† | Delta HDL (HDL baseline-HDL latest, mean ± SD, mg/dl) | p value† | Delta LDL (LDL baseline- LDL latest, mean ± SD, mg/dl) | p  value† |
| APOε2ε2 | No (n=12) | 11 ± 39 mg/dl | **7e-3** | 3 ± 10 mg/dl | 1e-1 | 12 ± 38 mg/dl | 2e-1 |
| APOε2ε2 | Yes (n=94) | 49 ± 57 mg/dl |  | -(2) ± 13 mg/dl |  | 28 ± 42 mg/dl |  |
| APOε2ε3 | No (n=335) | 9 ± 39 mg/dl | **5e-22** | -(2) ± 12 mg/dl | 7e-1 | 9 ± 30 mg/dl | **2e-20** |
| APOε2ε3 | Yes(n=1697) | 33 ± 47 mg/dl |  | -(2) ± 12 mg/dl |  | 27 ± 39 mg/dl |  |
| APOε3ε3 | No(n=1423) | 12 ± 39 mg/dl | **9e-74** | (0) ± 11 mg/dl | **6e-6** | 10 ± 32 mg/dl | **1e-85** |
| APOε3ε3 | Yes (n=9100) | 34 ± 48 mg/dl |  | -(2) ± 12 mg/dl |  | 30 ± 42 mg/dl |  |
| APOε2ε4 | No(n=69) | 12 ± 38 mg/dl | **3e-5** | -(2) ± 13 mg/dl | 9e-1 | 10 ± 35 mg/dl | **6e-5** |
| APOε2ε4 | Yes(n=306) | 35 ± 49 mg/dl |  | -(2) ± 13 mg/dl |  | 30 ± 41 mg/dl |  |
| APOε3ε4 | No(n=479) | 15 ± 1 mg/dl | **5e-20** | 0 ± 10 mg/dl | **2e-3** | 11 ± 34 mg/dl | **7e-26** |
| APOε3ε4 | Yes(n=3195) | 35 ± 50 mg/dl |  | -(2) ± 12 mg/dl |  | 30 ± 43 mg/dl |  |
| APOε4ε4 | No(n=43) | 15 ± 42 mg/dl | **1e-03** | 0 ± 10 mg/dl | 3e-1 | 12 ± 41 mg/dl | **1e-03** |
| APOε4ε4 | Yes(n=297) | 38 ± 52 mg/dl |  | -(2) ± 12 mg/dl |  | 35 ± 47 mg/dl |  |
| †Bold values having Bonferroni adjusted student t.test p value ≤0.004 (0.05/12) comparing delta lipids between non statin users and statin users.  Delta TC: A positive value suggest decrease in TC levels  Delta LDL: A positive value suggest decrease in LDL levels  Delta HDL: A negative value suggest increase in HDL levels  TC: Total cholesterol; HDL: High density lipoprotein; LDL: Low density lipoprotein; | | | | | | | |

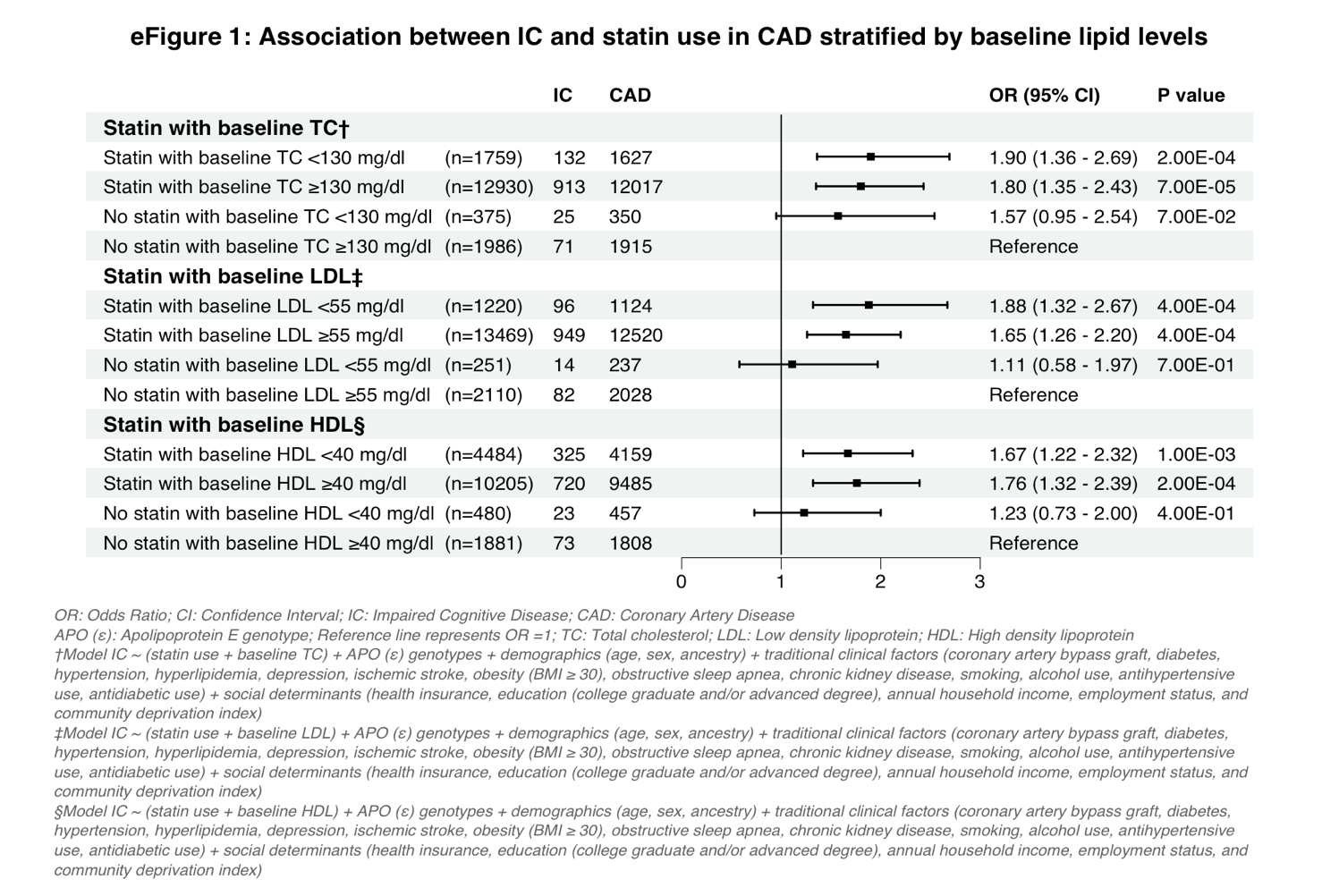

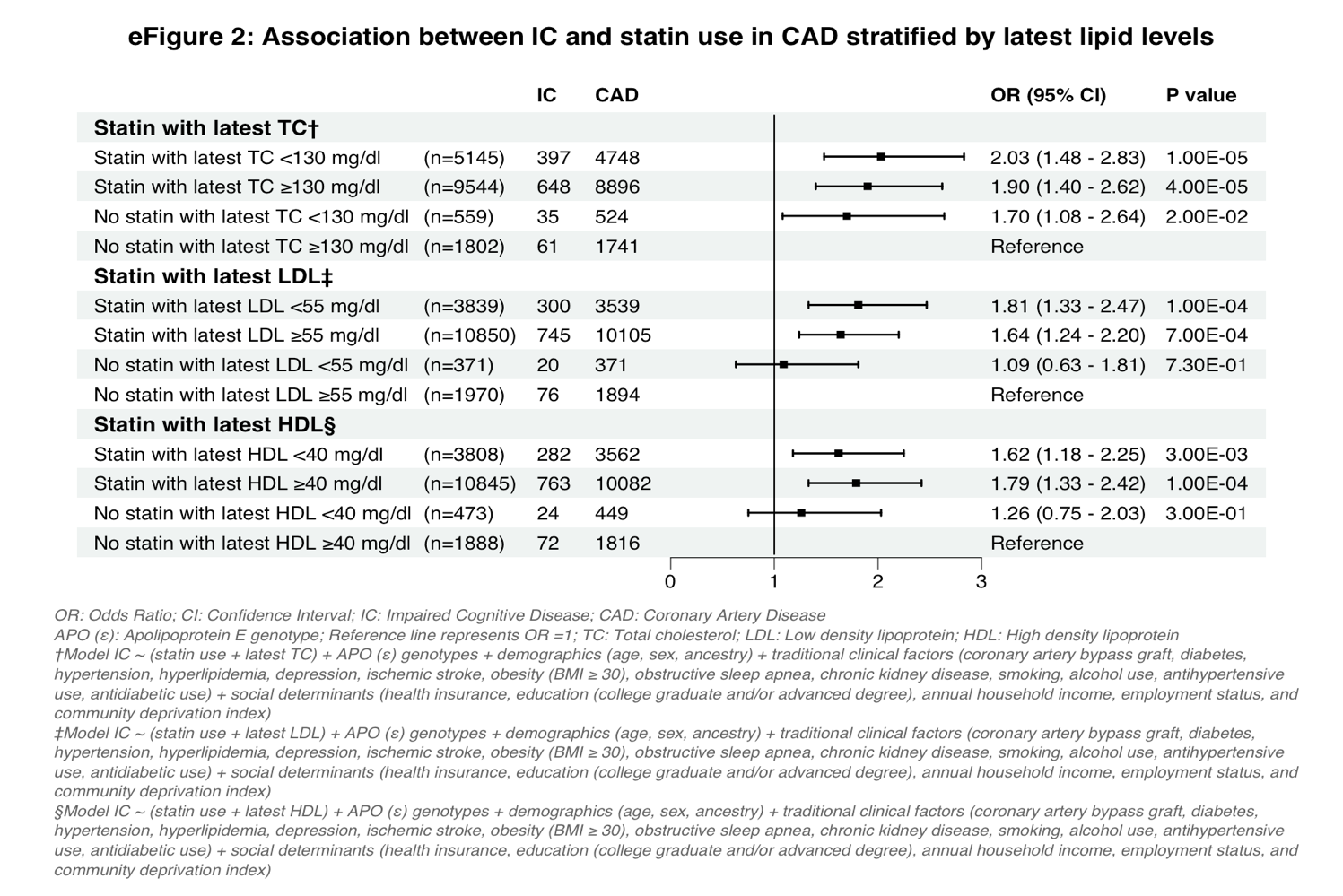

eFigure 3: Propensity matched plots in all CAD participants (IC with CAD & CAD without IC)

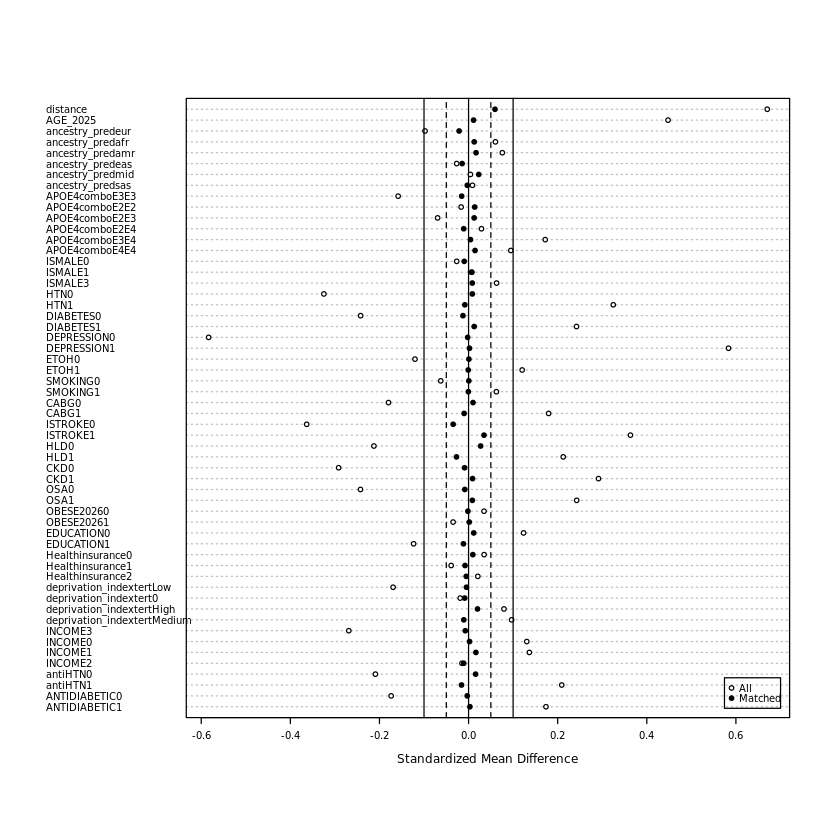

Notes:

ISMALE: Male sex (yes or no); HTN: Hypertension; ETOH: Alcohol use; HLD: Hyperlipidemia; ISTROKE: Ischemic stroke; OBESE2026: BMI ≥ 30; statin: antiHTN: antihypertensive drug use;CABG: Coronary artery bypass graft; APOE4combo: APO (ε) genotype; OSA: Obstructive sleep apnea; CKD: Chronic kidney disease; deprivation_indextert: Deprivation index tertile;AGE_2025: Age; EDUCATION: college graduate and/or advanced degree

CAD: Coronary Artery Disease

IC: Impaired Cognitive Disease

| Table 6: Baseline characteristics before and after propensity score matching (AllofUS, v8, Age ≥60) | | | | | | |
| --- | --- | --- | --- | --- | --- | --- |
| Before propensity score matching | | |  | **After propensity score matching** | | |
|  | CAD without IC (n=20,746) | IC with CAD (n=1,343) | P value | CAD without IC (n=4,029) | IC with CAD (n=1,343) | P value |
| Demographics* | | | | | | |
| Age (mean, SD) | 76.69 (62-105, SD=7.1) | 80.07 (64-105, SD=7.4) | **p<0.001** | 79.98 (63-105, SD=7.64) | 80.07 (64.0-105.0, SD=7.4) | p=0.73 |
| Female sex | 8,022 (38.7%) | 502 (37.4%) | p=0.36 | 1,525 (37.9%) | 502 (37.4%) | p=0.92 |
| Ancestry | | |  |  |  |  |
| Europeans | 16,024 (77.2%) | 979 (72.9%) | **p<0.001** | 2,975 (73.8%) | 979 (72.9%) | p=0.52 |
| Africans | 2,586 (12.5%) | 196 (14.6%) | **p=0.02** | 570 (14.1%) | 196 (14.6%) | p=0.72 |
| Admixed Americans | 1,738 (8.4%) | 144 (10.7%) | **p=0.003** | 411 (10.2%) | 144 (10.7%) | p=0.62 |
| Others | 398 (1.9%) | 24 (1.7%) | **p=0.82** | 73 (1.9%) | 24 (1.7%) | p=0.99 |
| Clinical factors | | | | | | |
| Depression | 7,731 (37.3%) | 874 (65.1%) | **p<0.001** | 2,940 (73.0%) | 874 (65.1%) | p=0.95 |
| Ischemic Stroke | 2,526 (12.2%) | 384 (28.6%) | **p<0.001** | 1,089 (27.0%) | 384 (28.6%) | p=0.26 |
| Hyperlipidemia | 19,330 (93.2%) | 1,301 (96.9%) | **p<0.001** | 3,922 (97.3%) | 1,301 (96.9%) | p=0.36 |
| Hypertension | 18,867 (90.9%) | 1,299 (96.7%) | **p<0.001** | 3,903 (96.9%) | 1,299 (96.7%) | p=0.78 |
| Diabetes | 9,924 (47.8%) | 802 (59.7%) | **p<0.001** | 2,381 (59.1%) | 802 (59.7%) | p=0.68 |
| Obesity | 9,161 (44.2%) | 570 (42.4%) | p=0.220 | 1,707 (42.4%) | 570 (42.4%) | p=0.96 |
| Highest Education | 10,086 (48.6%) | 571 (42.5%) | **p<0.001** | 1,736 (43.1%) | 571 (42.5%) | p=0.71 |
| Statin use | 16,019 (77.2%) | 1,172 (87.3%) | **p<0.001** | 3,382 (83.9%) | 1,172 (87.3%) | **p=0.003** |
| Antihypertensive use | 17,980 (86.7%) | 1,239 (92.3%) | **p<0.001** | 3,734 (92.7%) | 1,239 (92.3%) | p=0.61 |
| Antidiabetic use | 9796 (47.2%) | 750 (55.8) | **p<0.001** | 2,244 (55.7%) | 750 (55.8) | p=0.92 |
| CABG | 7,734 (37.3%) | 621 (46.2%) | **p<0.001** | 1,883 (46.7%) | 621 (46.2%) | p=0.75 |
| Obstructive sleep apnea | 7,556 (36.4%) | 652 (48.5%) | **p<0.001** | 652 (48.5%) | 652 (48.5%) | p=0.78 |
| Chronic kidney disease | 7,294 (35.2%) | 668 (49.7%) | **p<0.001** | 668 (49.7%) | 668 (49.7%) | p=0.77 |
| Social determinants | | | | | | |
| Health insurance | 20,141 (97.1%) | 1294 (96.4%) | p=0.248 | 3,888 (96.5%) | 1294 (96.4%) |  |
| Smoking | 4,734 (22.8%) | 343 (25.5%) | **p=0.022** | 1,030 (25.6%) | 343 (25.5%) | p=0.98 |
| Alcohol Use | 1,783 (8.6%) | 169 (12.6%) | **p<0.001** | 508 (12.6%) | 169 (12.6%) | p=0.98 |
| CDI (Low)† | 8,282 (39.9%) | 430 (32.0%) | **p<0.001** | 1,299 (32.2%) | 430 (32.0%) | p=0.92 |
| CDI (Medium/High)† | 11,780 (56.7%) | 873 (65%) | **p<0.001** | 2,604 (64.6%) | 873 (65%) | p=0.92 |
| Annual Income > $75,000 | 6,591 (31.8%) | 280 (20.8%) | **p<0.001** | 852 (21.1%) | 280 (20.8%) | p=0.95 |
| Employment (Retired) | 13,327 (64.2%) | 965 (71.9%) | **p<0.001** | 2,872 (71.3%) | 965 (71.9%) | p=0.86 |
| APO (ε) genotypes | | | | | | |
| APOε2ε2 | 133 (0.6%) | 7 (0.5%) | p=0.72 | 17 (0.4%) | 7 (0.5%) | p=0.81 |
| APOε2ε3 | 2,451 (11.8%) | 131 (9.8%) | **p=0.02** | 378 (9.4%) | 131 (9.8%) | p=0.73 |
| APOε3ε3 | 12,925 (62.3%) | 731 (54.4%) | **p<0.001** | 2,224 (55.2%) | 731 (54.4%) | p=0.64 |
| APOε2ε4 | 445 (2.1%) | 35 (2.6%) | p=0.30 | 112 (2.8%) | 35 (2.6%) | p=0.81 |
| APOε3ε4 | 4,404 (21.2%) | 390 (29.0%) | **p<0.001** | 1,162 (28.8%) | 390 (29.0%) | p=0.92 |
| APOε4ε4 | 388 (1.9%) | 49 (3.6%) | **p<0.001** | 136 (3.4%) | 49 (3.6%) | p=0.69 |
| *Proportions in bold with P value <0.05 (Pearson's Chi-squared test, baseline group = CAD) | | | | | | |
| Obesity (Body mass index ≥ 30) | | | | | | |
| Highest Education: College graduate and/or advanced degree | | | | | | |
| CAD: Coronary Artery Disease; IC: Impaired Cognitive Disease; CABG: Coronary Artery Bypass Graft | | | | | | |
| †CDI: Community Deprivation index. CDI not reported in (n=724) participants | | | | | | |

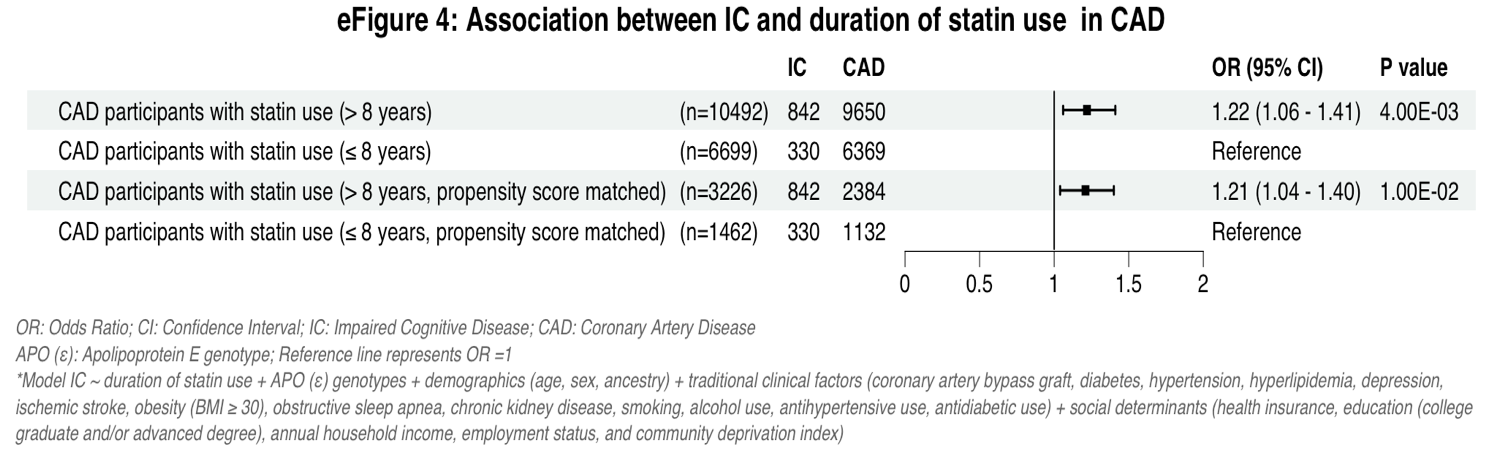

eFigure 5: Propensity matched plots in all CAD participants with statin use (IC with CAD & CAD without IC)

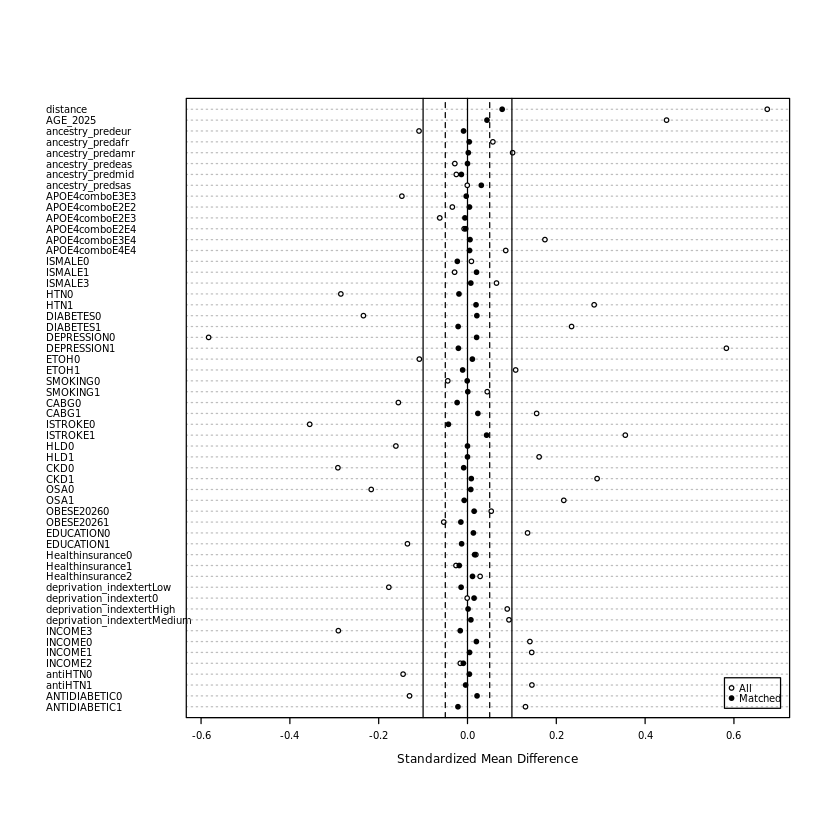

Notes:

ISMALE: Male sex (yes or no); HTN: Hypertension; ETOH: Alcohol use; HLD: Hyperlipidemia; ISTROKE: Ischemic stroke; OBESE2026: BMI ≥ 30; statin: antiHTN: antihypertensive drug use;CABG: Coronary artery bypass graft; APOE4combo: APO (ε) genotype; OSA: Obstructive sleep apnea; CKD: Chronic kidney disease; deprivation_indextert: Deprivation index tertile;AGE_2025: Age; EDUCATION: college graduate and/or advanced degree

CAD: Coronary Artery Disease

IC: Impaired Cognitive Disease

| Table 7: Exploring the Mind survey in statin users with CAD, AllofUS, v8, Age ≥60* | | | |
| --- | --- | --- | --- |
| Gradual Onset Continuous Performance Task:GradCPT (City or Mountain) | | | |
|  | Statin duration ≤ 8 yrs (n=317) | Statin duration > 8 yrs (n=495) | P Value |
| GradCPT: d-prime, Mean (SD, range) | 2.38 (0.77, 0.69-4.76) | **2.25 (0.72, 0.61-4.21)** | **p=0.004** |
| *Includes all coronary artery disease (CAD) participants with statin use who completed the optional GradCPT test in 2023  †Bold values with higher proportions having p value <0.01;  GradCPT assesses core executive function by testing sustained attention and inhibitory control.  d-prime: a measure of discrimination ability that represents the participant's ability to withhold responses to mountains while making responses to the more prevalent city images | | | |
